# Foundation models for generalizable electrocardiogram interpretation: comparison of supervised and self-supervised electrocardiogram foundation models

**DOI:** 10.1101/2025.03.02.25322575

**Authors:** Alexis Nolin-Lapalme, Achille Sowa, Jacques Delfrate, Olivier Tastet, Denis Corbin, Merve Kulbay, Derman Ozdemir, Marie-Jeanne Noël, François-Christophe Marois-Blanchet, François Harvey, Surbhi Sharma, Minhaj Ansari, I-Min Chiu, Valentina Dsouza, Sam F. Friedman, Michaël Chassé, Brian J. Potter, Jonathan Afilalo, Pierre Adil Elias, Gilbert Jabbour, Mourad Bahani, Marie-Pierre Dubé, Patrick M. Boyle, Neal A. Chatterjee, Joshua Barrios, Geoffrey H. Tison, David Ouyang, Mahnaz Maddah, Shaan Khurshid, Julia Cadrin-Tourigny, Rafik Tadros, Julie Hussin, Robert Avram

## Abstract

**Background:** The 12-lead electrocardiogram (ECG) remains a cornerstone of cardiac diagnostics, yet existing artificial intelligence (AI) solutions for automated interpretation often lack generalizability, remain closed-source, and are primarily trained using supervised learning, limiting their adaptability across diverse clinical settings. To address these challenges, we developed and compared two open-source foundational ECG models: DeepECG-SSL, a self-supervised learning model, and DeepECG-SL, a supervised learning model.

**Methods:** Both models were trained on over 1 million ECGs using a standardized preprocessing pipeline and automated free-text extraction from ECG reports to predict 77 cardiac conditions. DeepECG-SSL was pretrained using self-supervised contrastive learning and masked lead modeling. The models were evaluated on six multilingual private healthcare systems and four public datasets for ECG interpretation across 77 diagnostic categories. Fairness analyses assessed disparities in performance across age and sex groups, while also investigating fairness and resource utilization.

**Results:** DeepECG-SSL achieved AUROCs of 0.990 (95%CI 0.990, 0.990) on internal dataset, 0.981 (95%CI 0.981, 0.981) on external public datasets, and 0.983 (95%CI 0.983, 0.983) on external private datasets, while DeepECG-SL demonstrated AUROCs of 0.992 (95%CI 0.992, 0.992), 0.980 (95%CI 0.980, 0.980) and 0.983 (95%CI 0.983, 0.983) respectively. Fairness analyses revealed minimal disparities (true positive rate & false positive rate difference<0.010) across age and sex groups. Digital biomarker prediction (Long QT syndrome (LQTS) classification, 5-year atrial fibrillation prediction and left ventricular ejection fraction (LVEF) classification) with limited labeled data, DeepECG-SSL outperformed DeepECG-SL in predicting 5-year atrial fibrillation risk (N=132,050; AUROC 0.742 vs. 0.720; Δ=0.022; P<0.001), identifying reduced LVEF ≤40% (N=25,252; 0.928 vs. 0.900; Δ=0.028; P<0.001), and classifying LQTS syndrome subtypes (N=127; 0.931 vs. 0.853; Δ=0.078; P=0.026).

**Conclusion:** By releasing model weights, preprocessing tools, and validation code, we aim to support robust, data-efficient AI diagnostics across diverse clinical environments. This study establishes self-supervised learning as a promising paradigm for ECG analysis, particularly in settings with limited annotated data, enhancing accessibility, generalizability, and fairness in AI-driven cardiac diagnostics.

**Key Question:** Can self-supervised (SSL) learning yield ECG-based AI foundational models with enhanced performance, fairness, privacy, and generalizability compared to traditional supervised learning (SL) approaches?

**Key Finding:** Our evaluation of DeepECG-SL and DeepECG-SSL across seven external health center datasets and four international publicly accessible datasets demonstrated that while both models achieve comparable diagnostic accuracy for ECG interpretation, SSL outperforms SL on novel tasks with smaller datasets.

**Take-home Message:** We validated DeepECG-SL and DeepECG-SSL across public and private datasets and demonstrated that SSL model had a superior generalizability by addressing fairness, privacy, and efficiency, and open sourcing our models, we advance ethical, adaptable AI for equitable, real-world ECG diagnostics.

Graphical abstract:
DeepECG-SL and DeepECG-SSL, two open-source AI models for 12-lead ECG interpretation, were trained on over 1 million ECGs. DeepECG-SSL, utilizing self-supervised contrastive learning and masked lead modeling, outperformed DeepECG-SL in utilizing digital biomarkers to predict atrial fibrillation risk, reduced LVEF, and long QT syndrome subtypes, while both models achieved high diagnostic accuracy with minimal fairness disparities across age and sex. Validated on ten external datasets, our work provides a robust, reproducible framework for equitable, efficient ECG-based cardiac diagnostics.

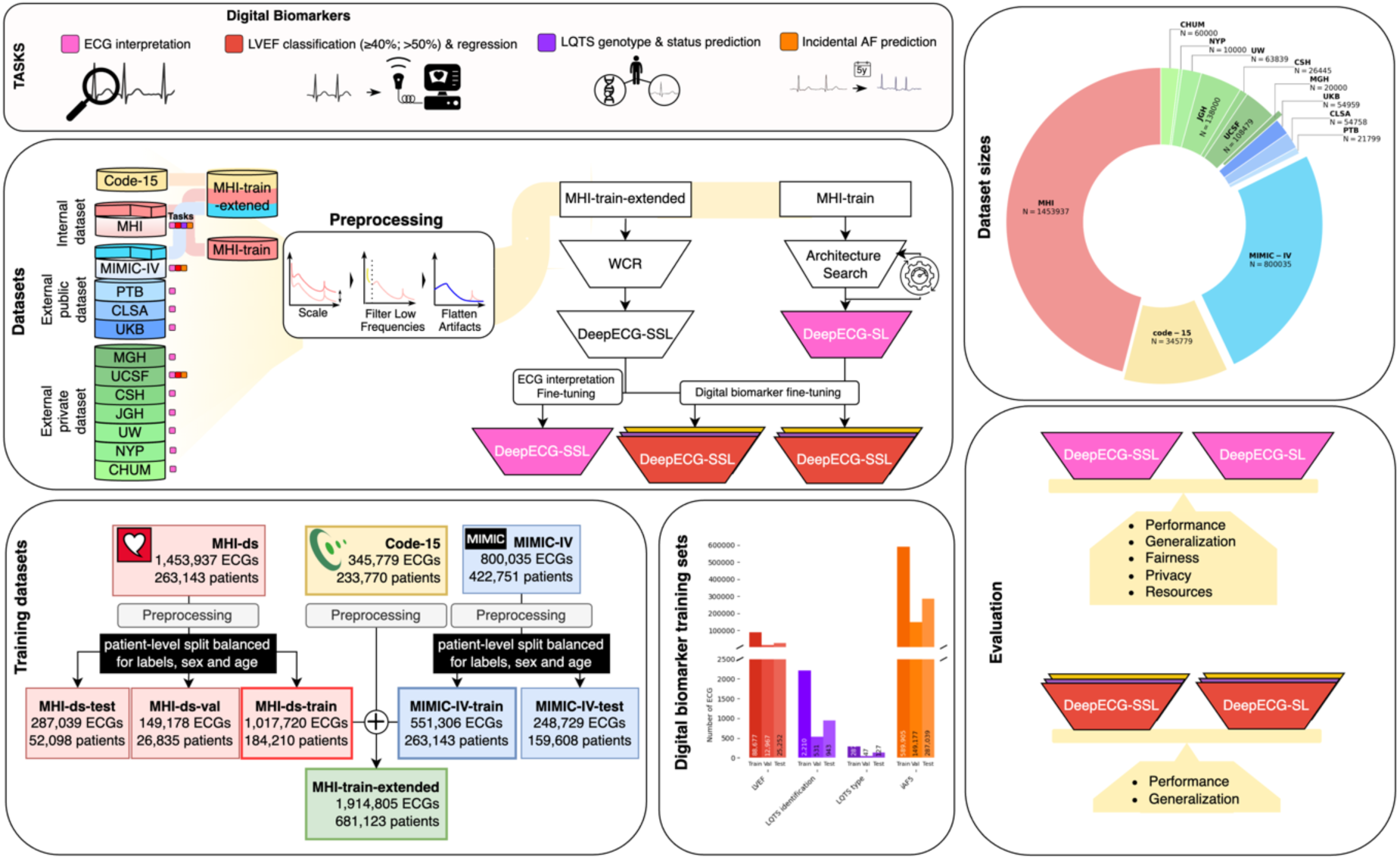

## Introduction

The 12-lead electrocardiogram (ECG) is a crucial diagnostic tool for cardiovascular disease, with over 300 million performed globally each year^1^. Beyond its traditional role in detecting arrhythmias and myocardial ischemia, where artificial intelligence (AI) algorithms can yield noteworthy performance approaching or exceeding expert clinician accuracy^2,3^, advances in ECG-AI analysis enable it to act as a digital biomarker of disease not typically diagnosed through ECG alone, such as presence of left ventricular dysfunction^4^, pulmonary hypertension^5^, and inherited arrhythmia genotype status, like long QT syndrome (LQTS)^6^ among others.

State-of-the-art models in ECG-AI analyses primarily rely on supervised learning (SL), which requires extensive labeled datasets tailored to specific, narrowly defined tasks^7^. In contrast, self-supervised learning (SSL) uses vast amounts of unlabeled data to pretrain models by discovering patterns and representations within the data itself^8^. These pretrained models can then be fine-tuned with smaller labeled datasets for specific tasks, often achieving superior performance and broader generalizability across diverse medical applications^9,10,11^. Despite these potential advantages of SSL, there are limited comparative studies evaluating both approaches for ECG-AI algorithms. Moreover, only 14% of medical AI^12^ research openly shares resources, thus significantly hindering reproducibility and collaborative progress^13^.

In this study, we present two ECG foundation models, large-scale models trained for multi-task generalizability: DeepECG-SL trained using a SL multilabel classification task and DeepECG-SSL pre-trained using self-supervised contrastive learning and masked lead modeling^14^ in over 1.9 million ECGs from three datasets. We then fine-tuned and evaluated the performance of both models to detect (i) 77 electrocardiographic diagnoses grouped into six categories (Supplementary Table 1) and digital biomarker tasks: (ii) echocardiographic left ventricular dysfunction and ejection fraction (LVEF), (iii) 5-year incidence of atrial fibrillation (iAF5), and (iv) LQTS diagnosis and genetic subtype. We evaluated our approach across eleven datasets (seven institutional and four public), encompassing 881,403 ECGs. This comprehensive validation assessed predictive performance, fairness, privacy, and computational efficiency.

## Methods

This retrospective study was approved by the Ethics Board of the Montreal Heart Institute (MHI), and all research activities were in accordance with the Declaration of Helsinki (2013)^38^. During manuscript preparation, large language models were used to assist with editorial and stylistic revisions. The authors take full responsibility for the scientific integrity, accuracy, and interpretation of the content presented herein.

### Training Datasets

The internal dataset, Montreal Heart Institute (MHI) dataset (*MHI-ds*), contains 1,453,937 12-lead ECGs from 263,277 patients. As a specialized quaternary care hospital, the MHI treats a wide range of cardiovascular conditions, making its dataset ideal for training a foundation model. These ECGs were recorded at the MHI between April 11, 1997, and November 10, 2023, using the MUSE system (General Electric, Boston), and were randomly split into training (*MHI-ds-train* - 70%), validation (*MHI-ds-val* - 10%), and test (*MHI-ds-test* - 20%) subsets at a patient level, ensuring that no single patient’s ECG appeared in more than one split. Age, sex, and label distributions were carefully balanced across all splits (Graphical abstract). *MHI-train-extended* dataset combined MHI-ds-train with two public datasets: the Code-15 dataset (345,779 ECGs from 233,770 patients)^17^ and MIMIC-IV-train, representing 70% (551,306 ECGs from 112,546 patients) of the full MIMIC-IV dataset^18^ (Graphical abstract). We applied similar constraints on age, sex and label distribution when splitting MIMIC-IV^18^ into MIMIC-IV-train (70%) and MIMIC-IV-test (30%).

### External Validation

To evaluate our models beyond the scope of the MIMIC-IV IV-test and MHI-ds-test sets, we defined all publicly available ECG-focused datasets or biobanks as “External Public Datasets” (EPD). These include the Canadian Longitudinal Study on Aging^22^ (CLSA)—a stratified sample of 51,338 Canadian adults aged 45–85 years—from which we used baseline (2011) ECGs (N = 29,427) and first follow-up ECGs (2015–2018; N = 25,185). We also incorporated data from the UK Biobank (UKB) ^20^ [Project #20168] collected during the first imaging visit in 2014 and a follow-up in 2019 (N = 54,978 ECGs), as well as the Physikalisch-Technische Bundesanstalt^21^ (PTB) dataset with PTB-XL annotations (N = 21,799).

To further assess clinical performance, we validated both models at multiple healthcare institutions—collectively designated as the “External Health Center” (EHC) datasets. It comprises ECGs from the following healthcare centers: the University of California, San Francisco (UCSF; 108,479 ECGs), Massachusetts General Hospital (MGH; 20,000 ECGs), Cedars-Sinai Hospital (CSH; 26,445 ECGs), Jewish General Hospital (JGH; 138,000 ECGs), the University of Washington Medical Center (UW; 63,839 ECGs), New York-Presbyterian Hospital (NYP; 10,000 ECGs), the Centre hospitalier de l’Université de Montréal (CHUM; 60,000 ECGs). Each institution’s data was curated according to the same criteria used for the training set and the external public datasets, ensuring uniform preprocessing and evaluation (Additional details on each cohort are provided in Supplementary Table 35).

### Data Preprocessing

To address the initial generalization challenges of DeepECG-SL, we compared the frequency power spectra of the MHI-ds and UKB datasets. This analysis revealed a significant enrichment of low frequencies below 1 Hz and prominent artifactual peaks at 50 Hz in the UKB dataset, corresponding to patient movement and the frequency of the British A/C current^39^ respectively. Similar peaks were observed in other datasets, such as PTB, though without the low-frequency enrichment (Supplementary Figure 1-A). Additionally, discrepancies in frequency amplitude were identified across most datasets analyzed in this study. To standardize the datasets and mitigate these issues, we developed an automated preprocessing pipeline. First, we applied a 1 Hz high-pass filter when the signal magnitude below 1 Hz exceeds one logarithmic unit relative to the 1–30 Hz range^40^. Second, artifacts, such as those caused by A/C-induced 60 Hz interference^41^, were detected when their amplitude exceeds two standard deviations above the local mean. These artifacts and their harmonics were flattened using a local LOESS curve^42^. Finally, amplitude was scaled to match the amplitude range of our internal dataset MHI-ds, resulting in a smooth power spectrum (Supplementary Figure 1-B). This standardized approach significantly improved cross-dataset generalization performance while preserving clinically relevant features in the 1-30 Hz range^40^ (Supplementary Table 9). We applied this pipeline on all datasets, except MHI-ds which served as reference.

### ECG Interpretation Task

We employed a systematic approach to ensure high-quality ECG labeling and consistency across all datasets. First, two experienced cardiologists (≥4 years of ECG interpretation experience) independently annotated 10,075 (4,200 from MHI-ds, 3,811 from MIMIC-IV, and 2,064 from UKB unique diagnostic statements paired to the ECG waveform, using a standardized framework of 77 labels spanning six clinical categories based on recommendations from the American Heart Association^43^. Each ECG annotated was selected as it contained one unique diagnostic strings captured in the remaining ECGs. Waveforms were reviewed in a standard 12-lead format, and any discrepancies were resolved by consensus with one additional cardiologist. If a condition appeared at any point in the 10-second recording, it was labeled as present at the record level. Inter-rater reliability was high, with Cohen’s kappa coefficients exceeding 0.80 for all diagnostic categories. As all unique diagnostic strings we captured by our annotation process the remaining MHI-ds, UKB and MIMIC-IV ECGs were annotated through the propagation of these 10,075 sentences alongside with their multi-label annotation to 160,129 unique paragraphs across the datasets.

To develop a robust multilingual diagnostic classifier, we expanded the aforementioned 10,075 expert-labeled sentences into 160,129 paragraphs. Further augmentation included 150,000 shuffled paragraph pairs (using LLAMA 3.1 70B^15^) and bidirectional French-English translation, yielding 640,518 total paragraph-label pairs. This expanded dataset was split (70/10/20% train/validation/test) to train a BERT-based classifier, ensuring accurate mapping of clinical statements to our 77-category diagnostic framework. Further details on the BERT classification methodology and predictions can be found in Supplementary Table 3 and Supplementary Methodology S1 and Supplementary Figure 11.

We integrated external datasets through three complementary methods:

1. **Direct Mapping** – Diagnostic paragraphs in MHI-ds, MIMIC-IV and UKBB were aligned to our 77-label framework via an existing dictionary.
2. **Manual Dictionary Conversion** – Applied to PTB, ensuring precise label alignment.
3. **Automated Classification** – Used a fine-tuned BERT model for CLSA and external health center (EHC) datasets, streamlining label assignment at scale.

Further details on the annotation procedures, including handling multilingual content, are provided in the Supplementary Methods.

### Digital Biomarker Tasks

To assess the model’s capability on novel tasks, we focused on LVEF prediction and classification, LQTS detection and type classification, and iAF5 prediction. These tasks were chosen because we had sufficiently large, annotated datasets for each across our external validation sites, and our team has previously published on these clinical outcomes, allowing direct comparison with previous work. Moreover, they showcase varying levels of supervision complexity, ranging from binary and multi-threshold classification to continuous-value regression. We ensured that each specialized task used the same MHI-train and MHI-test split to prevent any data leakage during fine-tuning and subsequently evaluated the resulting models in additional external datasets.

For both DeepECG-SL and DeepECG-SSL, we started with the model trained for ECG interpretation. To fine-tune the model, we unfroze all the layers and adjusted the classifier to have a single output for all tasks. We used the Adam optimizer with an initial learning rate of 0.01, along with binary focal loss set to alpha=0.25 and gamma=2.0 or mean square error for the LVEF regression task. To adjust the learning rate during training, we applied the Cosine Annealing Learning Rate scheduler with a maximum of 25 epochs. To prevent a selection bias, we used an early stop with a patience of 3 (Further details in Supplemental Methods S5).

#### LVEF Prediction

For LVEF-related tasks, we leveraged the datasets of UCSF, MIMIC-IV and MHI-ds. Each ECG was paired with the closest trans-thoracic echocardiography report recorded within a time window of −90 to +7 days of ECG recordings. In cases where multiple ECGs matched this criterion for the same patient, only the closest instance was selected. For the regression task, we used the LVEF values, while for classification tasks, we dichotomized LVEF values as < 50% and ≤ 40% to correspond to different severity thresholds for two distinct classification tasks.

#### LQTS Identification and Subtype Classification

For the LQTS tasks, we followed an approach like Jiang *et al.*^6^ leveraging the MHI-ds. Patients were labeled based on the presence of pathogenic or likely pathogenic variants in the KCNQ1 or KCNH2 genes, or a familial LQTS-causing genetic variant, which allowed us to differentiate between LQTS type 1 and type 2. Variants of uncertain significance (VUS) were reviewed by electrophysiologists specializing in inherited arrhythmias and classified as either strong VUS or weak VUS. Patients with strong VUS were labeled as LQTS cases, while those with weak VUS were used as controls for LQTS identification. Positive cases were further subdivided into type 1 and type 2 for subtype classification. This design allowed us to create both an LQTS identification task and a subtype classification task.

#### iAF5 Prediction

For iAF5 labeling, we followed the methodology outlined in our previous study^33^. ECGs from UCSF, MIMIC-IV and MHI-ds were included if they had valid metadata, waveform data, and were confirmed to be in sinus rhythm. Additional exclusion criteria included a proximity of less than 30 days to cardiac surgery or the presence of atrial flutter.

### Algorithm Development and Evaluation

#### DeepECG-SL

DeepECG-SL was trained on MHI-train-ds for ECG interpretation across 77 diagnoses, through an extensive sweep comparing different architectures, including families of transformers^15^, convolutional neural networks, and mamba^16^. The training also explored various data augmentation strategies, training environment parameters, initialization schemes, regularization techniques, and preprocessing strategies. To identify the optimal SL architecture, we utilized the Weights and Biases platform^44^, leveraging a Bayesian optimizer to maximize the Area Under the Precision-Recall Curve (AUPRC) and ensure hyper-parameter optimization (Supplementary Table 7, Supplemental Methods S3). Initialized with the weights optimized for ECG interpretation, DeepECG-SL was then fine-tuned for each digital biomarker prediction task.

#### DeepECG-SSL

We employed a two-stage training process for DeepECG-SSL: pre-training with unlabeled ECG data from MHI-train-extended dataset to learn general waveform features, followed by fine-tuning with labeled MHI-train data for ECG interpretation tasks.

For the pre-training stage, we experimented with several SSL approaches (Supplementary Methods S4) and opted for the framework introduced by Oh *et al.*^14^ (Supplementary Table 8). It combines two contrastive methods: Wav2Vec^19^, originally developed for self-supervised learning of speech representations and adapted here for learning local features, and Contrastive Multi-Segment Coding^45^, a method specifically designed for self-supervised learning of cardiac signals over time, for learning global representations. The inherent similarities between ECG and speech signals make this approach particularly well-suited for learning representations from raw ECG data (Supplemental Figure 2).

Both models were trained using four NVIDIA RTX A6000 graphic processing units (GPU)s. The architecture search for DeepECG-SL was conducted in parallel with one GPU for each model family, with each search running for two weeks. DeepECG-SSL was pre-trained on three NVIDIA RTX A6000 GPUs over a period of 14 days.

### Fairness and Privacy Evaluation

To ensure equitable performance, we assessed algorithmic fairness using the Equalized Odds framework^24^, which evaluates whether error rates are balanced across specific groups. In this analysis, we categorized age into three groups: under 55, 55–75, and over 75. Fairness was assessed by evaluating the performance of both models on the ECG interpretation task, where the TPR and FPR were computed for each group defined by age and sex, using micro-averaging. In each of the 1000 iterations, we randomly sampled 70% of the dataset and computed the TPR and FPR for each group. The average absolute disparity in TPR and FPR between groups was then calculated across all labels for each iteration. This process was repeated 1000 times to derive the mean disparity and 95% confidence intervals for both TPR and FPR differences. This methodology ensures a robust evaluation by incorporating sampling variability across multiple iterations.

For privacy evaluation, we applied MIA^25^. A Random Forest classifier with 50 trees was trained on the model logits to identify vulnerable predictions, informing privacy risk mitigation strategies. To estimate the model’s vulnerability with confidence intervals, the training process was repeated 10 times using a predefined set of environment seeds. In each iteration, the MHI-ds-train-derived logits and the testing logits were merged and appropriately labeled. The merged dataset was then split into 80% for training and 20% for testing to ensure a comprehensive evaluation of the model’s vulnerability detection capabilities.

To assess feature importance, we used the Random Forest classifier’s feature importance, averaging it across all 10 iterations. To visualize the logit distribution, we randomly sampled 10,000 entries from both the training and testing probability distributions, combining the two sets for visualization. Using the t-SNE algorithm with a perplexity of 30, we reduced the data to two dimensions for visualization. A scatter plot was then generated, with distinct colors representing the training and test samples, enabling a clear comparison of the logit distributions. This visualization provides insights into how well the model distinguishes between training and testing data in reduced-dimensional space.

### Explainability

To investigate the segments of the ECG signal that are key for a given prediction, we leveraged Local Interpretable Model-Agnostic Explanations^27^ (LIME). Briefly, LIME introduces localized perturbations to the signal and evaluates how these perturbations affect the output prediction. This approach enables the attribution of specific signal segments to the model’s decision-making process, providing valuable insights into the regions of the ECG that most influence predictions. Values present regions that positively influence the prediction. As the models were trained in a multilabel format, each analysis was conducted independently for each label associated with any given ECG. The analyses were performed in parallel for both DeepECG-SL and DeepECG-SSL, enabling a comparative assessment of the regions each model considers most relevant to its predictions.

### Evaluation Metrics and Statistical Analysis

We evaluated model performance using AUROC (for discrimination across thresholds), Area Under the Precision-Recall Curve (AUPRC; a better metric for performance in imbalanced classes), and sensitivity/specificity, with 95% confidence intervals derived through bootstrapping (70% data, 1,000 iterations) and AUROC comparisons using DeLong test^46^. Overall AUROC, representing global performance through concatenated label comparison, was used to compare the models. When confidence intervals were non-overlapping, we reported ΔSSL-SL (performance difference between models). To ensure optimal threshold for sensitivity and specificity, we optimized each label’s threshold to yield the best mean between sensitivity and specificity. These thresholds were computed on the MHI-ds-train and used on all the remaining datasets. For systematic evaluation, we categorized validation datasets into External Public Datasets (EPD: MIMIC-IV, PTB, CLSA, UKB) and External Healthcare Centers (EHC: UW, UCSF, MGH, CSH, NYP, JGH, CHUM), with detailed center-level performance metrics available in Supplementary Tables 11-24. We performed sensitivity analyses for the impact of the training set size on model performance, and we compared DeepECG-SSL against other SSL approaches such as Joint Embedding Predictive Architecture^47^ and **S**imple Framework for Contrastive Learning of Visual Representations^48^.

## Results

DeepECG-SL and DeepECG-SSL represent two distinct approaches to ECG-AI analysis. DeepECG-SL is trained on 1,017,720 ECGs across 184,210 patients. The models were developed using the MHI-ds-train, which maintains balanced demographics (mean age 60.40 years±0.08 5.87% male bias), with DeepECG-SSL additionally leveraging ECG signal data from Code-15 and MIMIC-IV datasets in the MHI-train-extended dataset for pre-training (Table 1). After an extensive sweep (670 different training runs), DeepECG-SL achieved the highest performance for ECG interpretation on MHI-ds-val using a modified EfficientNet-V2-S architecture^16^ optimized for one-dimensional signals (Supplementary Table 7, Supplementary Table 34). DeepECG-SSL employed a two-stage approach, with self-supervised pre-training and fine-tuning.

**Table 1:**
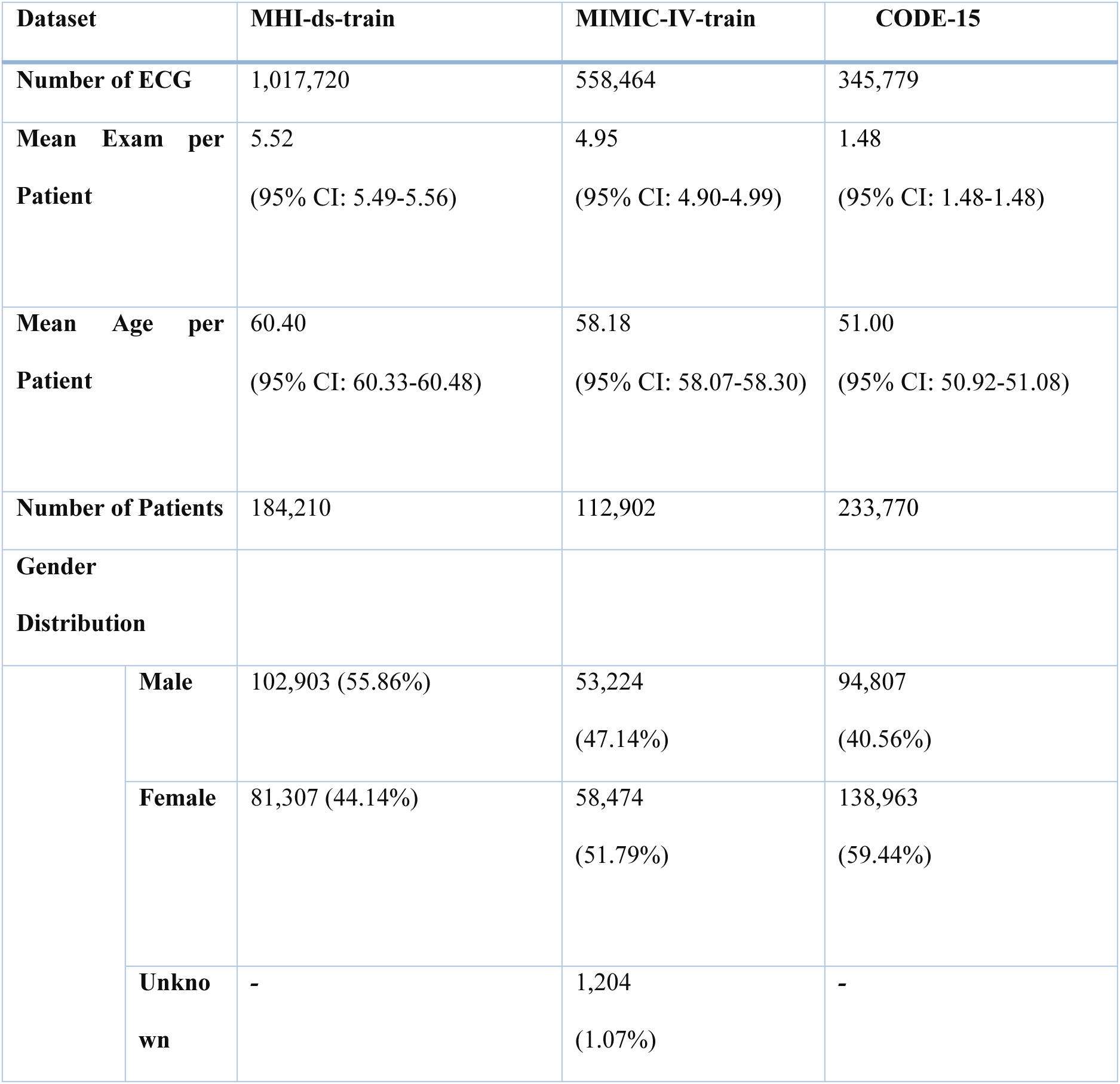
Overall demographics of the training set used in this study. . MHI-ds-train was used to train DeepECG-SL, and in combination with MIMIC-IV-train and Code-15 for DeepECG-SSL. To calculate the age distribution, the age was first averaged across all patients and then averaged throughout the dataset. The gender distribution is reported at the patient level. ***Abbreviations***: **MHI** (Montreal Heart Institute Dataset), **MIMIC-IV** (Medical Information Mart for Intensive Care IV Dataset), **ECG** (Electrocardiogram)

|  |  |  |  |
| --- | --- | --- | --- |
| Dataset | MHI-ds-train | MIMIC-IV-train | CODE-15 |

**Table 1: Overall demographics of the training set used in this study.**
|  |  |  |  |  |
| --- | --- | --- | --- | --- |
| **Number of ECG** |  | 1,017,720 | 558,464 | 345,779 |
| **Mean Exam per Patient** |  | 5.52<br>(95% CI: 5.49-5.56) | 4.95<br>(95% CI: 4.90-4.99) | 1.48<br>(95% CI: 1.48-1.48) |
| **Mean Age per Patient** |  | 60.40<br>(95% CI: 60.33-60.48) | 58.18<br>(95% CI: 58.07-58.30) | 51.00<br>(95% CI: 50.92-51.08) |
| **Number of Patients** |  | 184,210 | 112,902 | 233,770 |
| **Gender Distribution** |  |  |  |  |
|  | **Male** | 102,903 (55.86%) | 53,224<br>(47.14%) | 94,807<br>(40.56%) |
|  | **Female** | 81,307 (44.14%) | 58,474<br>(51.79%) | 138,963<br>(59.44%) |
|  | **Unknown** | - | 1,204<br>(1.07%) | - |

### ECG Interpretation Task

Our BERT annotation model achieved high classification performance, with the AUROC > 0.999 across all classes (except U wave, AUROC = 0.982) on the test set, which included 20% of the PTB, MIMIC-IV, and MHI-ds datasets. These predictions served as accurate ground truth for the EHC and CLSA datasets (Supplementary Table 2), whereas the other EPD underwent manual annotation.

Our comprehensive preprocessing pipeline, which transforms raw ECG signals through frequency-domain cleaning (including 1Hz high-pass filtering, power spectrum normalization, and artifact removal), significantly improved model performance across datasets (Δ AUROC up to 0.251; MHI-ds: 0.992 vs. 0.741, CLSA: 0.995 vs. 0.828, UKB: 0.988 vs. 0.813), while maintaining high performance on already-preprocessed datasets (PTB and MIMIC-IV) (Supplementary Table 9).

Both models demonstrated consistently high performance across the internal MHI-ds test set and a broad spectrum of external datasets spanning public repositories (EPD) and private health centers (EHC), as illustrated in Figure 2 and Supplementary Tables 11–23. On the MHI internal test set (N=156,721), DeepECG-SL achieved an AUROC of 0.992 (95% CI: 0.992-0.992) and AUPRC of 0.912, slightly outperforming DeepECG-SSL (AUROC: 0.989; AUPRC: 0.909). Across external public datasets (EPD; N=131,517), both models demonstrated robust generalizability with comparable performance (DeepECG-SL AUROC: 0.981, AUPRC: 0.897; DeepECG-SSL AUROC: 0.980, AUPRC: 0.895). Similarly, on external healthcare center datasets (EHC; N=545,539), both models maintained high performance (AUROC: 0.983 for both; AUPRC: 0.901 vs 0.900).

Performance remained consistent across major diagnostic categories including rhythm disorders (AUROC >0.92), conduction abnormalities (AUROC >0.96), and chamber enlargement (AUROC >0.92), with statistically significant but clinically minimal differences between models (all Δ <0.01). Notable exceptions included acute and chronic infarct and ischemia detection tasks on external public datasets, where both models showed reduced performance for infarct and ischemia detection (DeepECG-SL AUROC: 0.867 vs DeepECG-SSL: 0.863; P<0.001) compared to MHI (0.984 vs 0.980; P<0.001) and external healthcare centers (0.954 vs 0.954; P=0.583). This pattern suggests that ischemic detection may be more challenging in public datasets, which typically include fewer acute cases compared to healthcare settings. Detailed performance metrics across all diagnostic categories and datasets are provided in Supplementary Tables 11-24.

### Digital Biomarker Tasks

We evaluated three AI-derived digital biomarkers - ECG-based predictions that extend beyond standard electrocardiographic interpretation. These included: (1) LQTS detection and subtype classification (KCNQ1 vs KCNH2 variants), (2) left ventricular ejection fraction estimation (classification tasks: LVEF ≤40% and <50% and regression tasks), and (3) 5-year *de novo* atrial fibrillation risk prediction from sinus rhythm ECGs (iAF5). These tasks, having limited labeled data availability, served as stringent tests of model generalization across healthcare institutions. For LQTS, based on Jiang et al’s^6^ method we subdivided it into two classification tasks: the prediction of LQTS, defined as ECGs from individuals with a pathogenic or likely pathogenic variant in the *KCNQ1* or *KCNH2* genes, and the subtask of LQTS subtype distinction, distinguishing between KCNQ1- and KCNH2-based types. These labels were solely available in the MHI-ds (for more details see <u>Methods</u>).

Performance of both models on internal MHI-ds is presented using AUROC (Figure 1C) and AUPRC (Supplementary Table 25). On digital biomarker tasks, DeepECG-SSL outperformed DeepECG-SL in predicting 5-year atrial fibrillation risk (AUROC: 0.742 vs 0.720; Δ=0.022; P<0.001) and identifying reduced ejection fraction ≤40% (AUROC: 0.928 vs 0.900; Δ=0.028; P<0.001), while maintaining comparable performance for LVEF <50 and LQTS detection. For external validation, both models were assessed on the EPD and EHC datasets (Figure 1-C, Supplementary Table 25-33). DeepECG-SSL consistently delivered superior performance for iAF5, LVEF ≤ 40, and LVEF < 50 across these diverse institutions and populations (all *P* < 0.001, except for LVEF < 50 in the University of Washington dataset).

**Figure 1.**
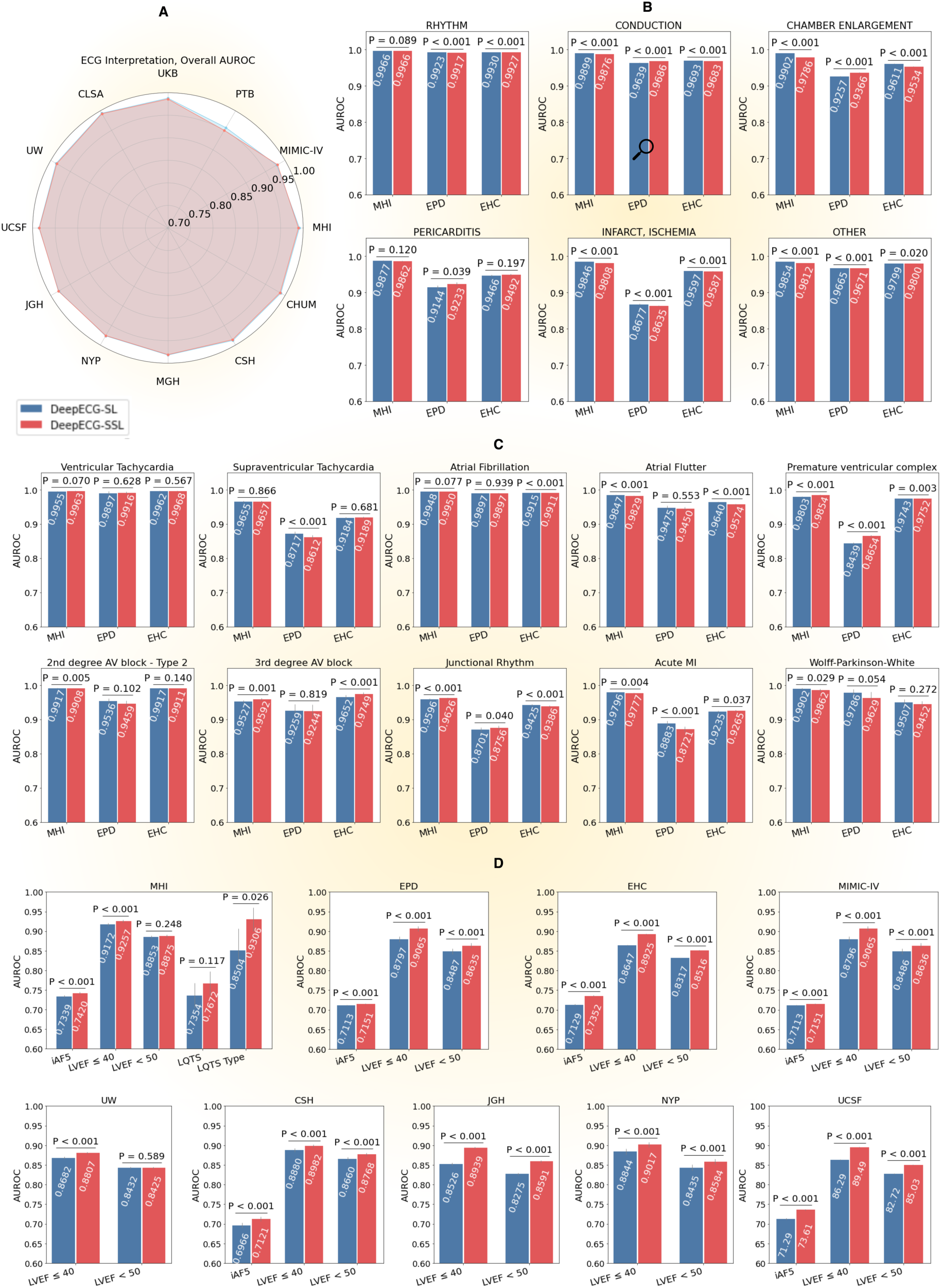
An overview of DeepECG-SL and DeepECG-SSL performances. We report overall AUROC and P-value computed using the DeLong Test. **A** Overall AUROC of ECG interpretation across all datasets. **B** AUROC of ECG interpretation categories on MHI-ds, EPD and EHC. **C**. AUROC of ECG interpretation categories on MHI-ds, EPD and EHC. **D** AUROC of digital biomarkers across datasets. iAF5 is at ECG level **Abbreviations**: **CSH** (Cedars Sinai Hospital Dataset), **CLSA** (Canadian Longitudinal Study on Aging Dataset), **ECG** (Electrocardiogram), **EHC** (External Health Centers Dataset), **EPD** (External Public Dataset), **JGH** (Jewish General Hospital Dataset), **LQTS** (Long QT Syndrome), **LVEF** (Left Ventricular Ejection Fraction), **MGH** (Massachusetts General Hospital Dataset), **MHI** (Montreal Heart Institute Dataset), **MIMIC-IV** (Medical Information Mart for Intensive Care IV Dataset), **NYP** (New York-Presbyterian Hospital Dataset), **PTB** (Physikalisch-Technische Bundesanstalt Dataset), **UKB** (UK Biobank Dataset), **UCSF** (University of California San Francisco Medical Center Dataset), **UW** (University of Washington Medical Center), **CHUM** (Centre hospitalier de l’Université de Montréal), and **WCR** (Wave2Vec + Contrastive Multi-segment Coding + Random Lead Masking).

We analyzed how dataset size influences model performance (Supplemental Figure 4). For large datasets (ECG interpretation, N=1,166,896), both models performed similarly. For moderately-sized tasks (iAF5 detection, N=532,742), the performance gap was minimal (AUROC: 0.742 vs 0.734). However, for tasks with limited data (LQTS-Type classification, N=334), DeepECG-SSL significantly outperformed DeepECG-SL (AUROC: 0.931 vs 0.850; Δ=0.081; P=0.026).

To further validate this relationship, we conducted experiments using varying percentages of the MHI training data. While both models maintained comparable performance when using ≥10% of the data, DeepECG-SSL showed significant advantages when training data was severely limited (1% of original dataset), suggesting particular utility for rare conditions or emerging clinical applications.

### Fairness and Privacy

*Fairness* is described as ensuring that AI systems provide equitable outcomes across diverse groups, minimizing biases and promoting equal treatment. One metric to quantify fairness is equalized odds^17^, which ensures that the model’s true positive rate (TPR) and false positive rate (FPR) are similar across different groups, thereby reducing disparate impact. TPR, also known as sensitivity, measures the proportion of actual positives correctly identified by the model, while FPR captures the proportion of negatives incorrectly classified as positives. Thus, a smaller TPR and FPR difference across groups demonstrates higher rates of fairness. Both models demonstrated strong fairness across demographics (gender-based TPR/FPR differences <0.01), with DeepECG-SSL showing marginally better balance across age and gender groups (Supplementary Table 4).

Privacy evaluation through membership inference attacks (MIA)^18^ involves assessing whether an adversary can determine if a specific data point was used to train a machine learning model using the models’ output logits. By evaluating its vulnerability to such attacks, it is possible to gauge the privacy risks of deploying the model and, more importantly, identify the factors that lead to distinguishing features between training and testing samples. Both models showed strong privacy preservation on internal MHI-ds data (AUROC <0.6) and PTB dataset (AUROC <0.78). However, external datasets with distinct feature distributions, particularly MIMIC-IV and UKB, showed higher re-identification rates (AUROC >0.95), suggesting domain shifts rather than model architecture drive privacy vulnerabilities (Supplementary Table 5).

Analysis of membership inference attack patterns revealed that re-identification success was driven by dataset characteristics rather than model architecture. Datasets where models relied heavily on specific features showed higher re-identification rates, while those with distributed feature importance demonstrated better privacy protection. T-SNE visualization^19^ of model outputs confirmed this pattern: greater separation between training and test set representations correlated with higher re-identification rates (Supplementary Figures 5-8). For instance, in CLSA, DeepECG-SL’s disproportionate weighting of specific features (“No QRS,” “Regular,” “Monomorph”) led to more distinct clusters and increased re-identification risk.

### Resource Usage

DeepECG-SSL demonstrates higher computational demands compared to DeepECG-SL across several metrics. The SSL model is substantially larger (90.37M vs 1.51M parameters; 60-fold increase) and requires more operations (14.17 GMAC vs 530.57 MMAC; 27-fold increase). For processing 1000 ECGs on GPU, DeepECG-SSL consumes more energy (0.7463 vs 0.1786 Wh; 318% increase) and produces higher CO₂ emissions (1.774 vs 0.425 mgCO₂). To contextualize, processing 1 million ECGs with DeepECG-SSL produces CO₂ emissions equivalent to driving a car for 9.6 seconds at 100mph, compared to 17 seconds for DeepECG-SL (Supplemental Table 6, Supplemental Methods 6).

### Explainability

To understand the model’s decision-making process, we employed Local Interpretable Model-Agnostic Explanation^20^ (LIME) on a subset of representative ECG signals. Analysis demonstrates that both DeepECG-SSL and DeepECG-SL focus on clinically relevant features (Supplemental figure 9,10) demonstrating that both supervised and self-supervised learning methods yield valuable feature learning. However, when compared to the same examples, the intensity and selectivity of the pattern differs with DeepECG-SSL appearing to rely on fewer regions of the ECG than DeepECG-SL. This suggests that DeepECG-SSL may learn more concentrated or discriminative features, potentially enabling more targeted predictions without compromising overall classification performance.

## Discussion

In this study, we introduce two foundation models: DeepECG-SL and DeepECG-SSL. Both rigorously evaluated on four critical clinical tasks: ECG interpretation (encompassing 77 diagnoses), LVEF prediction, 5-year incident atrial fibrillation prediction, and LQTS detection with subtype classification. Both models achieved state-of-the-art performance on our internal dataset and demonstrated robust generalizability across ten geographically diverse external datasets (373,865 ECGs in four public datasets and 447,538 ECGs in six private healthcare centers) totaling 821,403 ECGs. In this comprehensive validation across 10 medical centers and 821,403 ECGs—one of the largest in ECG AI—both models demonstrated robust clinical performance, maintaining AUROC >0.90 for 76% of diagnostic categories despite diverse patient populations and institutional labeling practices. Our findings demonstrate distinct clinical advantages for each model: DeepECG-SSL excelled in detecting emerging biomarkers with limited training data (LVEF classification, LQTS detection), while the more efficient DeepECG-SL (60-fold smaller, 29-fold faster inference) matched performance on traditional ECG diagnoses. Both models showed minimal performance degradation across external validations (AUROC decline 0.02-0.05), supporting their reliability across diverse clinical settings. To facilitate clinical adoption, we provide comprehensive open-source access to model weights, preprocessing tools, multilingual diagnostic extraction, and validation code. This standardized framework facilitates cost-effective local fine-tuning and reproducible implementation.

Recent advances in ECG foundation models includes ECGFounder^21^, a supervised algorithm trained on 10 million ECGs, ECG-FM^22^ a self-supervised model, and KED^23^ (Knowledged-enhanced ECG) a contrastive learning-based model. All have demonstrated progress in automated ECG interpretation. ECGFounder achieved high internal performance (AUROC ≥ 0.95 in 82 out of 150 diagnostic labels) but required an order of magnitude more data than our study. ECG-FM, which shares our WCR-based^14^ SSL training method, SSL training strategy, employed 5-second ECG segments to predict 13 diagnostic labels—doubling the training data but omitting temporally dependent patterns vital for detecting conditions such as second-degree AV blocks. The KED framework used contrastive learning to align ECG signals with ECG text reports. It demonstrated impressive diagnostic performance on single-label classification tasks, with AUROC often exceeding 0.90, without requiring additional training on multiple external datasets. used contrastive learning to align ECG signals with text reports, enabling potentially infinite textual labels; however, it is primarily suited to single-label tasks, may struggle with multi-label scenarios, and did not evaluate or release its model for conditions not explicitly mentioned in ECG reports (e.g., left ventricular dysfunction). DeepECG qualifies as a foundation model because it is trained on a large, heterogeneous dataset encompassing a broad range of clinical conditions, contains an extensive number of parameters, and demonstrates strong performance across multiple external datasets. Moreover, it exhibits label-efficient adaptability, making it suitable for various downstream tasks without full re-training. This aligns with the Stanford HAI definition^24^, which emphasizes comprehensive pretraining on broad data to enable general-purpose capabilities.

Our models demonstrated consistently high AUROC (> 0.95) for common diagnoses such as sinus rhythm, atrial fibrillation, right bundle branch block, and ventricular pacing. Performance remained robust even for more complex labels like Wolff-Parkinson-White, though it varied in rarer conditions (such as Brugada), where limited sample sizes likely contributed to wider confidence intervals. Notably, digital biomarker predictions such as LVEF, 5-year incident atrial fibrillation risk, and LQTS—traditionally requiring echocardiography or genetic testing—were accurately derived from routine ECGs, highlighting the potential for earlier, non-invasive detection in clinical practice. For instance, both DeepECG-SL and DeepECG-SSL surpassed Hou et al.’s^25^ ECG-based LVEF < 50 prediction benchmark, while also delivering lower mean absolute errors (MAE) in LVEF regression. Similarly, both models improved upon our team’s prior iAF5^26^ detection model (mean AUROC difference: 0.015 at ECG-level) on the same dataset we previously reported for validation. On LQTS detection, Jiang et al.^6^ reported marginally higher AUROCs on MHI-ds, but the differences from our model were not statistically significant (overlapping confidence intervals). Their model was trained on a unique Canadian registry (4,521 ECGs), whereas ours used a smaller MHI-ds subset (2,741 LQTS ECGs), limiting direct comparability. Access to their full training data might further improve our performance.

We addressed four critical barriers to real-world AI adoption—fairness, privacy, resource efficiency, and explainability—linking technical rigor to clinical trust. Fairness evaluations revealed minimal disparities in TPR and FPR across age and sex, mitigating biases against underdiagnosed populations. DeepECG-SSL’s strong performance across diverse centers suggests self-supervised learning can reduce label-related biases. To gauge privacy risks, we used MIA and t-SNE clustering, showing how specialized centers like MHI could be re-identified due to distinct disease distributions. Safeguards such as differential privacy^27^ may reduce this risk but can slightly impact AUROC (drops of 0.03–0.05). Resource analysis confirmed DeepECG-SL’s suitability for low-resource environments, with faster inference, while DeepECG-SSL’s larger size excelled in data-scarce tasks, even on CPU-based setups. Finally, explainability via LIME showed both models rely on clinically meaningful waveform features, with DeepECG-SSL focusing on narrower segments—an approach akin to expert cardiologists’ attention. This transparency supports trustworthy integration into routine ECG workflows, augmenting rather than disrupting clinical practice. In terms of workflow integration, we envision running these models alongside traditional ECG systems to complement rather than replace existing interpretations. This “dual reporting” approach can bolster diagnostic confidence, refine triage priorities, and ultimately reduce unnecessary downstream testing and is currently being tested^28^ in a randomized controlled trial (NCT 06462989).

Our open-source foundation model targets a critical unmet need by improving diagnostic access in resource-limited settings. Traditional ECG-AI solutions often require large, site-specific labeled data or proprietary software^29^, which hinders practical deployment in smaller hospitals or under-resourced regions. In contrast, DeepECG’s label-efficient training and robust performance—even for low-data digital biomarker tasks—allow rapid local adaptation with minimal computational overhead. Notably, no comparable model is readily available for straightforward container-based deployment: our Docker solution can output predictions in standard EMR systems with minimal additional infrastructure. By enabling earlier detection of conditions like LQTS or low LVEF—often underdiagnosed where advanced cardiac testing is scarce—this approach holds promise for more equitable and cost-effective cardiovascular care worldwide.

Despite promising performance, several critical limitations warrant consideration. While UK Biobank results suggest minimal demographic bias, our predominantly North American validation cohorts necessitate broader evaluation across diverse populations, particularly regarding racial, socioeconomic, and comorbidity dimensions. The proposed “dual-reporting” approach requires careful examination of clinical workflow integration, including protocols for AI-human discrepancies, alert fatigue mitigation, and liability considerations. Although LIME-based explanations provide interpretability, their clinical utility and alignment with established ECG criteria need rigorous validation through structured cardiologist feedback. Label variability in EHR-dependent datasets remains a concern, though robust external performance partially mitigates this issue. Moreover, we did not directly compare model performance with an expert committee, but our reported accuracy aligns with or surpasses that of prior AI studies tested against cardiologist-level performance^30^. Our framework’s demonstrated efficiency— achieving AUROC 0.97 with just 100,000 ECGs—suggests sufficient performance with modest dataset sizes, contrasting with more data-intensive domains. However, practical implementation faces substantial hurdles, including regulatory approval pathways, EMR integration costs, and computational resource requirements. Future work should prioritize prospective validation studies, systematic failure mode analysis (particularly for rare pathologies), and exploration of multimodal data integration, while maintaining focus on real-world clinical utility. While our accuracy aligns with or exceeds previous AI studies, direct comparison with expert committees would provide valuable validation of clinical readiness. Lastly, all p-values do not take into account multiple testing.

We present two extensively validated ECG foundation models across multiple clinical settings. DeepECG-SSL excels in emerging biomarker detection with limited training data (including LQTS subtyping and LVEF classification), while the more efficient DeepECG-SL matches performance on traditional diagnoses despite being substantially smaller. Both models demonstrate robust external generalization across most diagnostic categories, minimal demographic bias, and clinically aligned feature detection. Our open-source pipeline enables reproducible deployment and efficient local adaptation through standardized preprocessing and Dockerized implementation. An ongoing randomized trial evaluates clinical integration alongside existing ECG systems, with future work focused on expert committee validation and expanded clinical applications. These contributions establish a comprehensive framework for responsible, clinically validated AI in cardiovascular diagnostics

## Data availability

The internal MHI dataset and external private datasets from health centers UW, UCSF, MGH, JGH, CSH, and NYP are not publicly available due to its potentially identifiable nature of patients in these datasets. However, MIMIC-IV is accessible from https://physionet.org/content/mimiciv/3.1/, UKB is available upon request from https://www.ukbiobank.ac.uk/enable-your-research/apply-for-access, Code-15 can be accessed from https://zenodo.org/records/4916206, PTB is available from https://physionet.org/content/ptb-xl/1.0.3/, and CLSA is available upon request from https://www.clsa-elcv.ca/data-access/.

## Code availability

The code for data preprocessing, model training, and fine-tuning is available on the GitHub page: https://github.com/HeartWise-AI/DeepECG_Docker/tree/main. The model weights can also be accessed from the same page.

## Acknowledgments

We would like to thank Maxime Belanger from the IT department at MHI for his invaluable assistance with deployment and support in managing the computer infrastructure. Additionally, we extend our gratitude to Matthew Scicluna for his insightful contributions to the t-SNE visualization and his assistance with the analysis. We also thank Dr Sushravya Raghunath for his great help with the NYP dataset.

## Author Contribution

**Alexis Nolin-Lapalme** – Assisted with ECG annotation, developed the analysis pipeline, developed DeepECG-SL and wrote the manuscript.

**Achille Sowa** – Developed the DeepECG-SSL pipeline and developed the analysis pipeline and wrote the manuscript.

**Jacques Delfrate** – Developed the Docker container and assisted with the deployment on EHC. Contributed to the analysis, assisted with the revision of the manuscript.

**Olivier Tastet** – Assisted with the analysis and assisted with the revision of the manuscript.

**Denis Corbin** – Assisted with the analysis and assisted with the revision of the manuscript.

**Merve Kulbay, Derman Ozdemir, & Marie-France Noël** – Assisted with ECG annotation and assisted with the revision of the manuscript.

**Surbhi Sharma, Patrick M. Boyle, & Neal A. Chatterjee** – Conducted the UW validation and assisted with the revision of the manuscript.

**Minhaj Ansari & Joshua Barrios, Geoffrey H. Tison** – Conducted the UCSF validation, manuscript revision and assisted with the revision of the manuscript.

**I-Min Chiu & David Ouyang** – Conducted the CSH validation and assisted with the revision of the manuscript.

**Valentina D’Souza, Sam Friedman, Mahnaz Maddah, & Shaan Khurshid** – Conducted the MGH validation and assisted with the revision of the manuscript.

**Michael Chasse, Brian Potter, François-Christophe Marois-Blanchet, François Harvey** – Conducted the CHUM validation and assisted with the revision of the manuscript.

**Jonathan Afilalo** – Conducted the JGH validation and assisted with the revision of the manuscript.

**Pierre Adil Elias** – Conducted the NYP validation and assisted with the revision of the manuscript.

**Gilbert Jabbour** – Assisted with the iAF5 validation and assisted with the revision of the manuscript.

**Mourad Bahani** – Assisted with SSL methods and assisted with the revision of the manuscript.

**Marie-Pierre Dubé** – Assisted with the UKB and assisted with the revision of the manuscript.

**Julia Cadrin-Tourigny** – Contributed to the interpretation of the results and assisted with the revision of the manuscript.

**Rafik Tadros** – Contributed to the interpretation of the results and assisted with the revision of the manuscript.

**Julie Hussin** – Funding acquisition, supervision and contributed to the interpretation of the results and assisted with the revision of the manuscript.

**Robert Avram** – Funding acquisition, supervision, conceptualization, and manuscript writing and assisted with the revision of the manuscript.

## Glossary

BERT: bidirectional encoder representations from transformers
CHUM: Centre hospitalier de l’Université de Montréal
CLSA: Canadian Longitudinal Study on Aging
CSH: Cedars Sinai Hospital
EHC: external private health center datasets (UCSF, UW, NYP, JGH, MGH, CSH, CHUM)
EPD: external public datasets (CLSA, MIMIC-IV, PTB, UKB)
ECG: electrocardiogram
FPR: false positive rate
GMAC: Giga Multiply-Accumulate Operations
GPU: Graphic processing unit
iAF5: incident atrial fibrillation at 5 years
JGH: Jewish General Hospital
LQTS: Long-QT syndrome
LVEF: left ventricular ejection fraction
MAE: mean absolute error
MIA: membership inference attacks
MIMIC: Medical Information Mart for Intensive Care
MGH: Massachusetts General Hospital
MMAC: Mega Multiply-Accumulate Operations
MHI: Montreal Heart Institute
MHI-ds: Montreal Heart Institute Dataset
NYP: New York-Presbyterian Hospital
PTB: Physikalisch-Technische Bundesanstalt
RLM: Representation Learning with Masking
SL: supervised learning
SSL: self-supervised learning
t-SNE: t-distributed stochastic neighbor embedding
TPR: true positive rate
UCSF: University of California San Francisco Medical Center
UKB: UK Biobank
UW: University of Washington Medical Center
VUS: variants of uncertain significance
WCR: Wav2Vec + Contrastive Multi-segment Coding + Random Lead Masking

## References

1. Holst, H., Ohlsson, M., Peterson, C. & Edenbrandt, L. A confident decision support system for interpreting electrocardiograms. Clin. Physiol. 19, 410–418 (1999).

2. Martínez-Sellés, M. & Marina-Breysse, M. Current and Future Use of Artificial Intelligence in Electrocardiography. JCDD 10, 175 (2023).

3. Hannun, A. Y. et al. Cardiologist-level arrhythmia detection and classification in ambulatory electrocardiograms using a deep neural network. Nat Med 25, 65–69 (2019).

4. Yao, X. et al. Artificial intelligence–enabled electrocardiograms for identification of patients with low ejection fraction: a pragmatic, randomized clinical trial. Nat Med 27, 815–819 (2021).

5. Aras, M. A. et al. Electrocardiogram Detection of Pulmonary Hypertension Using Deep Learning. J. Card. Fail. 29, 1017–1028 (2023).

6. Jiang, R. et al. Deep Learning–Augmented ECG Analysis for Screening and Genotype Prediction of Congenital Long QT Syndrome. JAMA Cardiol 9, 377 (2024).

7. LeCun, Y., Bengio, Y. & Hinton, G. Deep learning. Nature 521, 436–444 (2015).

8. Krishnan, R., Rajpurkar, P. & Topol, E. J. Self-supervised learning in medicine and healthcare. *Nat*. Biomed. Eng. 6, 1346–1352 (2022).

9. Lai, J. et al. Practical intelligent diagnostic algorithm for wearable 12-lead ECG via self-supervised learning on large-scale dataset. Nat Commun 14, 3741 (2023).

10. Mehari, T. & Strodthoff, N. Self-supervised representation learning from 12-lead ECG data. Comput. Biol. Med. 141, 105114 (2022).

11. Zhou, Y. et al. A foundation model for generalizable disease detection from retinal images. Nature 622, 156–163 (2023).

12. Kelly, B. S. et al. Correction to: Radiology artificial intelligence: a systematic review and evaluation of methods (RAISE). Eur Radiol 32, 8054–8054 (2022).

13. Pineau, J., et al. Improving Reproducibility in Machine Learning Research (A Report from the NeurIPS 2019 Reproducibility Program). (2020) doi:10.48550/ARXIV.2003.12206.

14. Oh, J., Chung, H., Kwon, J., Hong, D. & Choi, E. Lead-agnostic Self-supervised Learning for Local and Global Representations of Electrocardiogram. (2022) doi:10.48550/ARXIV.2203.06889.

15. Grattafiori, A., et al. The Llama 3 Herd of Models. Preprint at 10.48550/ARXIV.2407.21783 (2024).

16. Tan, M. & Le, Q. V. EfficientNetV2: Smaller Models and Faster Training. (2021) doi:10.48550/ARXIV.2104.00298.

17. Hardt, M., Price, E. & Srebro, N. Equality of Opportunity in Supervised Learning. (2016) doi:10.48550/ARXIV.1610.02413.

18. Shokri, R., Stronati, M., Song, C. & Shmatikov, V. Membership Inference Attacks against Machine Learning Models. (2016) doi:10.48550/ARXIV.1610.05820.

19. Maaten, L. van der & Hinton, G. Visualizing Data using t-SNE. J. Mach. Learn. Res. 9, 2579–2605 (2008).

20. Ribeiro, M. T., Singh, S. & Guestrin, C. ‘Why Should I Trust You?’: Explaining the Predictions of Any Classifier. (2016) doi:10.48550/ARXIV.1602.04938.

21. Li, J., et al. An Electrocardiogram Foundation Model Built on over 10 Million Recordings with External Evaluation across Multiple Domains. (2024) doi:10.48550/ARXIV.2410.04133.

22. McKeen, K., et al. ECG-FM: An Open Electrocardiogram Foundation Model. (2024) doi:10.48550/ARXIV.2408.05178.

23. Tian, Y. et al. Foundation model of ECG diagnosis: Diagnostics and explanations of any form and rhythm on ECG. Cell Rep. Med. 5, 101875 (2024).

24. Casusi, J. What is a Foundation Model? An Explainer for Non-Experts. (2024).

25. Hou, Y. et al. Deep Learning-based 12-Lead Electrocardiogram for Low Left Ventricular Ejection Fraction Detection in Patients. Can. J. Cardiol. S0828282X24009802 (2024) doi:10.1016/j.cjca.2024.09.018.

26. Jabbour, G. et al. Prediction of incident atrial fibrillation using deep learning, clinical models, and polygenic scores. Eur. Heart J. 45, 4920–4934 (2024).

27. Dwork, C. Differential Privacy. in Automata, Languages and Programming (eds. Bugliesi, M., Preneel, B., Sassone, V. & Wegener, I.) vol. 4052 1–12 (Springer Berlin Heidelberg, Berlin, Heidelberg, 2006).

28. Avram, R. Harnessing ECG Artificial Intelligence for Rapid Treatment and Accurate Interpretation (HEART-AI). https://clinicaltrials.gov/study/NCT06462989.

29. Anumana AI., A. unlocking the language of the heart. Available.

30. Hughes, J. W. et al. Performance of a Convolutional Neural Network and Explainability Technique for 12-Lead Electrocardiogram Interpretation. JAMA Cardiol 6, 1285 (2021).

