## Supplementary material for "Foundation models for generalizable electrocardiogram interpretation: comparison of supervised and self-supervised electrocardiogram foundation models": Supp Figures and Methods

### Supplemental Methods

#### S1. Data annotation and BERT Training

To train the BERT-based model[1], we aggregated diagnostic paragraphs from expert-curated ECG interpretations across three datasets: MHI-ds, MIMIC-IV, and UKBB. The original reports were validated by cardiologists. Special characters were removed, and unique sentences were extracted across the datasets. We then created an ontology of 77 unique diagnostic classes, based on recommendations from Kligfield et al.[2], to serve as ground truth.

To standardize annotations, we identified 10,075 unique sentences (4,200 from MHI-ds, 3,811 from MIMIC-IV, and 2,064 from UKBB) and uploaded two examples of each to Labelbox.com for expert annotation. Two independent reviewers, each with ≥4 years of ECG interpretation experience, annotated these sentences, mapping them to predefined diagnostic categories, including chamber abnormalities, rhythm disorders, ischemic changes, and conduction abnormalities. For example, the sentence "Sinus bradycardia with 1st degree AV block with occasional premature" was annotated as ["Sinus Rhythm", "Regular", "Bradycardia", "Monomorph", "1st degree AV block"]. Discrepancies between annotations were adjudicated by a senior cardiologist with ≥10 years of experience to ensure consensus. Inter-rater reliability was high, with Cohen's kappa coefficients exceeding 0.80 for all diagnostic categories. Annotations were applied at the record level, meaning that if a condition (e.g., premature ventricular contraction) appeared even once during a 10-second ECG, the diagnosis was added. Consensus labels were then propagated to identical sentences across the three datasets. For non-unique sentences, labels were generated using a fine-tuned BERT model. The labeled datasets were combined and split into training (70%), validation (10%), and test (20%) sets.

The BERT-base-uncased architecture was fine-tuned on 640,518 multilingual (English/French) diagnostic paragraph-label pairs from UKBB, MHI-ds, and MIMIC-IV. The model training employed the focal loss function (gamma=2, alpha=0.25)[3], to address class imbalance, the AdamW[4] optimizer, a linear learning rate schedule (1e-6), and a batch size of 16. Model performance was evaluated on manually annotated test sets (MHI-ds and MIMIC-IV), demonstrating high prediction accuracy (see Supplement Table 2). This validated the model's effectiveness in automating label propagation for non-annotated data.

#### S2. Detailed Preprocessing Algorithm

All data formats including XML, WFDB, HDF5 or DICOM are first parsed to yield a NumPy matrix containing the 12 leads of the ECG in the following order: I, II, III, aVR, aVL, aVF, V1, V2, V3, V4, V5, V6 at 250Hz. The diagnostic text is also extracted and prepared in a list object containing the paragraphs as string for the BERT model.

To ensure uniformity across all datasets, we designed a preprocessing pipeline that is composed of a sequence of steps:

1. ECG signals were transformed into the frequency domain using Fast Fourier Transform (FFT)[5], enabling the analysis of signal power across various frequencies. The magnitude spectrum S(f) was calculated as the absolute value of the FFT output:

$$S(f) = |FFT(x)|$$

   Where x is the ECG signal in the time domain and f represents the corresponding frequencies. The FFT was applied individually to each signal in each lead in the dataset, and the resulting magnitude spectrum was used for further processing.

2. We applied a 1Hz high-pass filter to the ECG signal to remove low-frequency noise, such as baseline drift. This prevents distortion when components below 1Hz are more than twice the magnitude of those at 5Hz, ensuring clearer data by mitigating low-frequency enrichment caused by patient movement.

3. To ensure equivalent amplitude scaling in mV across all datasets, we compared the dataset's power spectrum between 1-30Hz with a reference curve, allowing us to yield an adjustment factor

$$factor = target\ power / mean\ spectral\ power$$

   The frequency components of each signal were scaled by this factor.

$$Adjusted\ Signal(t) = Signal(t) * factor$$

4. To detect and remove strong artifact peaks, such as those induced at 50 or 60Hz A/C peaks, a sliding window approach was used to scan the magnitude spectrum for prominent peaks. Each window's mean and standard deviation of magnitudes were computed, and peaks were identified as frequencies where the magnitude exceeded a threshold defined as:

$$Threshold = mean + k\ x\ std$$

where k is a user-defined constant. Detected peaks were further analyzed for harmonic relationships, defined as frequencies that were integer multiples (2x, 3x, 4x, etc.) of previously identified peaks. A tolerance for harmonic deviations was set at 5%, and harmonics were identified using a relative frequency threshold.

Once the peaks were detected, the signal was smoothed around these peaks. The LOESS (Locally Weighted Scatterplot Smoothing) method[6] was used to assume the flattened region to which the peak is flatten to.

5. The ECGs were regenerated using the Inverse Fast Fourier Transform (IFFT) returning the signal in the time domain.

$$y(t) = IFFT(Y(f))$$

#### S3. DeepECG-SL training

To train the DeepECG-SL model, we utilized the Weights and Biases API[7] for efficient experiment tracking and optimization. To comprehensively explore the full parameter space (Supplementary Table 7), we employed a Bayesian optimizer, which allowed extensive tuning of all possible parameters up to 500 iterations. We trained each model family on a separate GPU for seven days, ensuring that every family: ViT[8], CrossViT[9], EfficientNet[10],[11], Mamba[12], ResNet[13], ResNeXt[14], Inception[15], DenseNet[16] was afforded a complete exploration of its parameter space. Since the loss function itself was a parameter to be optimized, we focused on the validation AUPRC, given its challenging nature in multilabel contexts[17].

***Data Scaling Strategies***

For data scaling, the optimizer had the option to either leave the data unchanged or apply a variety of transformations, such as Min-Max Scaling, Max-Absolute Scaling, Quantile Scaling, Robust Scaling, and the Yeo-Johnson Power Transform[18]. These transformations could be applied on a per-dataset, per-sample, or per-lead basis, depending on the model's needs.

***Data Augmentation Strategies***

Regarding data augmentation, the optimizer could determine both the quantity and the type of random augmentations to apply in each batch. Augmentation strategies included techniques like window-warp-multithreaded, window-slice-multithreaded, time-warp-multithreaded, magnitude-warp-uniform-multithreaded, beat-permutation, scaling, jitter, or no augmentation at all. These methods were inspired by Iwana & Uchida[19], but were rewritten using JIT[20] to adapt 12-lead ECG signals and improve processing speed.

***Loss Functions***

The complexity of multilabel learning, combined with the dataset's imbalance, led the optimizer to explore various loss functions. These included TwoWayLoss[21], Hill Loss[22], Asymmetric Loss[23], SPLC[22], MultiLabelSoftMarginLoss, Binary Cross-Entropy (binary-ce), Dice Loss, Binary Focal Loss (gamma=2), Binary Focal Loss (gamma=3)[3], and Weighted Binary Cross-Entropy (weighted-bce). These loss functions were selected to optimize model performance in this challenging context.

***Activation Functions***

For model flexibility, we allowed the optimizer to select the activation function for each convolutional block and/or transformer block. Available activation functions included ReLU[24], Leaky ReLU[25], GELU[26], SELU[27], Mish[28], and Swish[29], providing fine-grained control over the model's nonlinearities.

***Stochastic Depth and Dropout***

The optimizer was able to adjust the stochastic depth[30], a technique where certain layers are randomly skipped during training to reduce overfitting. This, alongside adjustable dropout rates[31], allowed the optimizer to control model complexity and regularization more effectively.

***Training Settings***

We provided a range of options for the choice of training optimizer, including Adam[32], AdamW[4], Radam[33], SGD[34], Adagrad[35], and RMSprop[36]. The optimizer could also select an initial learning rate from a predefined range (0.01 to 0.000000001) and choose from different learning rate schedulers, including by-plateau, cosine-annealing[37], triangular-2, and lambda, among others. The optimizer could also apply a warmup phase, use an Exponential Moving Average (EMA) for model parameters, and employ adaptive gradient clipping[38]. Additionally, the optimizer was allowed to choose from various batch sizes, including 218, 512, 780, or 1024.

#### S4. DeepECG-SSL pre-training

We explored several self-supervised learning (SSL) methodologies for pre-training and assessed their performance on ECG interpretation. Specifically, we tested SIMCLR[39] and BYOL[40] with a ResNet-50[13] architecture, JEPA[41] with a transformer architecture, and WCR[42] with a CNN + Transformer architecture. The results of these experiments on ECG interpretation are summarized in Supplementary Table 1. Due to constraints in computational resources and the extensive time required to pre-train each model, we were unable to exhaustively explore all potential architectures and hyperparameter configurations. Ultimately, we selected WCR as it demonstrated the best performance on the MHI internal test set. We pre-trained DeepECG-SSL using WCR. As stated in Figure 5, it combines two contrastive losses, one local contrastive loss within each segment (half ECG block) and one global contrastive loss between segments. We used a learning rate of 5e-5, a batch size of 1026 (342 per GPU) and over 250 epochs. All other settings were kept at their default value.

**Supplementary Table 1: AUROC of different SSL approaches for ECG interpretation, on MHI internal dataset**

| Methods | Metrics | WCR (95% CI) | JEPA (95% CI) | SIMCLR (95% CI) | BYOL (95% CI) |
|---|---|---|---|---|---|
| RHYTHM | AUROC | **0.997 (0.997,0.997)** | 0.995 (0.995, 0.995) | 0.993 (0.993, 0.993) | 0.992 (0.992, 0.992) |
| | AUPRC | **0.985 (0.984,0.985)** | 0.979 (0.979, 0.979) | 0.967 (0.966, 0.967) | 0.965 (0.965, 0.965) |
| CONDUCTION | AUROC | **0.988 (0.988,0.988)** | 0.984 (0.983, 0.984) | **0.988 (0.987, 0.988)** | 0.986 (0.986, 0.987) |
| | AUPRC | **0.843 (0.842,0.844)** | 0.811 (0.810, 0.812) | **0.850 (0.849, 0.851)** | 0.838 (0.837, 0.839) |
| CHAMBER ENLARGEMENT | AUROC | 0.979 (0.978,0.979) | 0.986 (0.985, 0.986) | **0.989 (0.989, 0.990)** | **0.989 (0.989, 0.989)** |
| | AUPRC | 0.731 (0.728,0.733) | 0.842 (0.840, 0.844) | **0.883 (0.881, 0.884)** | **0.876 (0.874, 0.877)** |
| PERICARDITIS | AUROC | 0.986 (0.985,0.988) | 0.982 (0.980, 0.983) | **0.989 (0.989, 0.990)** | 0.986 (0.985, 0.987) |
| | AUPRC | 0.480 (0.465,0.493) | 0.469 (0.455, 0.483) | **0.520 (0.506, 0.534)** | 0.500 (0.485, 0.515) |
| INFARCT, ISCHEMIA | AUROC | 0.981 (0.981,0.981) | 0.975 (0.975, 0.976) | **0.983 (0.982, 0.983)** | 0.980 (0.980, 0.980) |
| | AUPRC | 0.719 (0.717,0.721) | 0.667 (0.665, 0.670) | **0.747 (0.744, 0.749)** | 0.729 (0.727, 0.731) |
| OTHER | AUROC | **0.981 (0.981,0.981)** | 0.979 (0.979, 0.980) | 0.981 (0.981, 0.981) | 0.979 (0.979, 0.979) |
| | AUPRC | **0.885 (0.885,0.886)** | 0.878 (0.877, 0.879) | 0.887 (0.887, 0.888) | 0.877 (0.877, 0.878) |
| OVERALL | AUROC | **0.990 (0.990,0.990)** | 0.988 (0.988, 0.988) | 0.988 (0.988, 0.988) | 0.987 (0.987, 0.987) |
| | AUPRC | **0.923 (0.922,0.923)** | 0.913 (0.913, 0.914) | 0.917 (0.917, 0.918) | 0.911 (0.910, 0.911) |

***Abbreviations***: BYOL (Bootstrap Your Own Latent), JEPA (Joint Embedding Predictive Architecture), SIMCLR (Simple Framework for Contrastive Learning of Visual Representations), WCR (Wave2Vec+Contrastive Multi-Segment Coding+Random Lead Masking)

#### S5 DeepECG-SSL and DeepECG-SL fine-tuning

We fine-tuned DeepECG-SSL and DeepECG-SL separately for each task. Each downstream model was initialized with the pretraining weights and fine-tuned using the corresponding subset of MHI-ds-train specific to the task. For DeepECG-SSL the architecture for each model comprised DeepECG-SSL encoder (pretrained model) and a classifier head consisting of a single-layer projection with number of output neurons corresponding to the number of labels (77 for ECG interpretation and 1 for the other tasks). In the case of DeepECG-SL the model was initialized with the best weights where only the output neurons were changed similarly to DeepECG-SSL. All the weights were updated during fine-tuning. Various loss functions were tested, and for external validation, we selected the loss function that achieved the best AUROC on the internal test set. For ECG interpretation, LVEF $\leq 40$ and LVEF $< 50$, we used binary cross-entropy loss. For iAF5, LQTS, and LQTS-Type, binary focal loss provided the best results. Finally for LVEF regression, we used mean squared error loss. All fine-tuning was conducted with a batch size of 128, a learning rate of 5e-5, the Adam optimizer, and trained for a maximum of 100 epochs.

#### S6 Power consumption and Model size analysis

We assessed the inference performance of both DeepECG-SL and DeepECG-SSL models on both GPU and CPU by running 100 repeated tests on a randomly sampled subset of 1000 examples. Inference time, $CO_2$ emissions, and energy consumption are tracked for each run using the CodeCarbon package[43]. The evaluation is performed separately for GPU and CPU, with results presented as the mean and 95% confidence intervals for each metric. To ensure reproducibility, a small batch size was used, allowing for a fair comparison across different hardware. The statistics for each device are reported, including inference time, $CO_2$ emissions, and energy consumption.

To evaluate the size of both models, we leveraged the Calflops package[44], which allows us to measure not only the number of parameters but also the number of operations, both in terms of MAC and FLOPs.

The average passenger vehicle emits about 400 grams of $CO_2$ per mile[45], which is equivalent to 400,000 milligrams of $CO_2$ per mile. To understand this emission in the context of GPU processing, we first calculate the time it takes to drive 1 mile at 100 miles per hour. Driving at this speed, it takes 36 seconds to cover 1 mile. Given that the car emits 11,111.11 milligrams of $CO_2$ per second (i.e., 400,000 mg per 36 seconds), we can now use this emission rate to compare the $CO_2$ emissions from processing 1000 examples.

We applied the following rule of three (proportional calculation) to relate GPU emissions to the car emissions:

$$\text{Emissions(seconds)} = \frac{\text{CO emissions(mg)}}{11,111.11\,\text{mg CO per second}}$$

#### S7 Explainability Analysis

To analyze the key ECG segments influencing label predictions, we utilized the Lime-For-Time wrapper built on Local Interpretable Model-Agnostic Explanations[46] (LIME). To achieve an optimal balance between computational efficiency and result quality, the 12x2500 ECG signal was divided into 1200 segments. Each segment was randomly perturbed by replacing its values with samples drawn from a uniform distribution bounded by the minimum and maximum values of the signal. This process was repeated for 1000 samples to ensure robust estimation. The resulting LIME values were mapped back onto the signal to generate the visualization. For clarity, only positive contributions were displayed, with negative values set to zero. Additionally, to simplify interpretation, the plots focused on the most clinically meaningful leads chosen by a cardiologist.

### Supplementary Figures

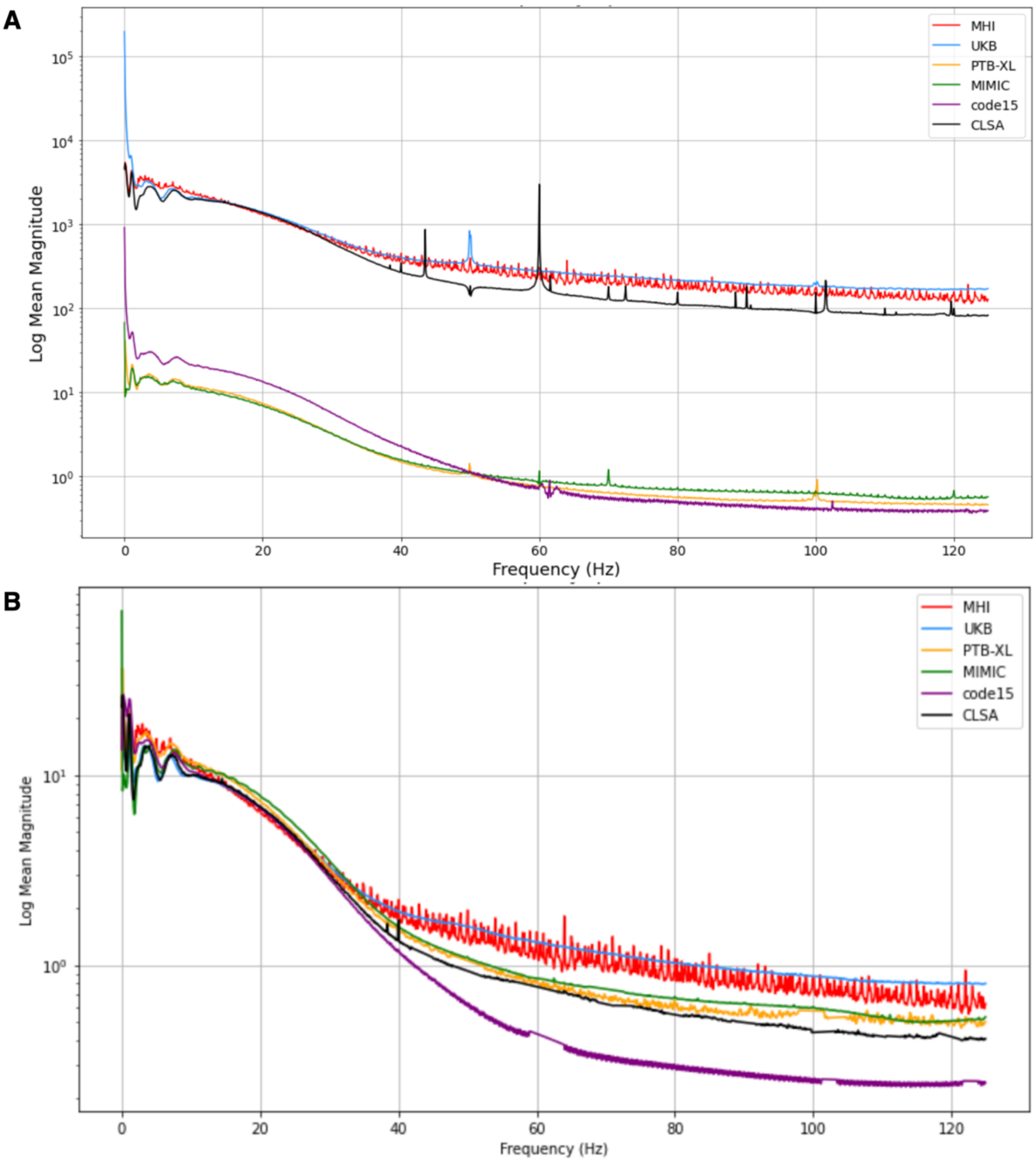


**Supplementary Figure 1: Power spectra of MHI-ds and EPD before and after preprocessing. A** Power spectrum of raw MHI-ds and EPD before any preprocessing is performed. **B** Power spectra following preprocessing of MHI-ds and EPD.

***Abbreviations***: **MHI-ds:** Montreal Heart Institute dataset, **EPD**: External Public Dataset (CLSA, UKB, PTB, MIMIC-IV), , **CLSA:** Canadian Longitudinal Study on Aging , **UKB:** UK Biobank, **PTB**: Physikalisch-Technische Bundesanstalt, **MIMIC-IV**: Medical Information Mart for Intensive Care IV

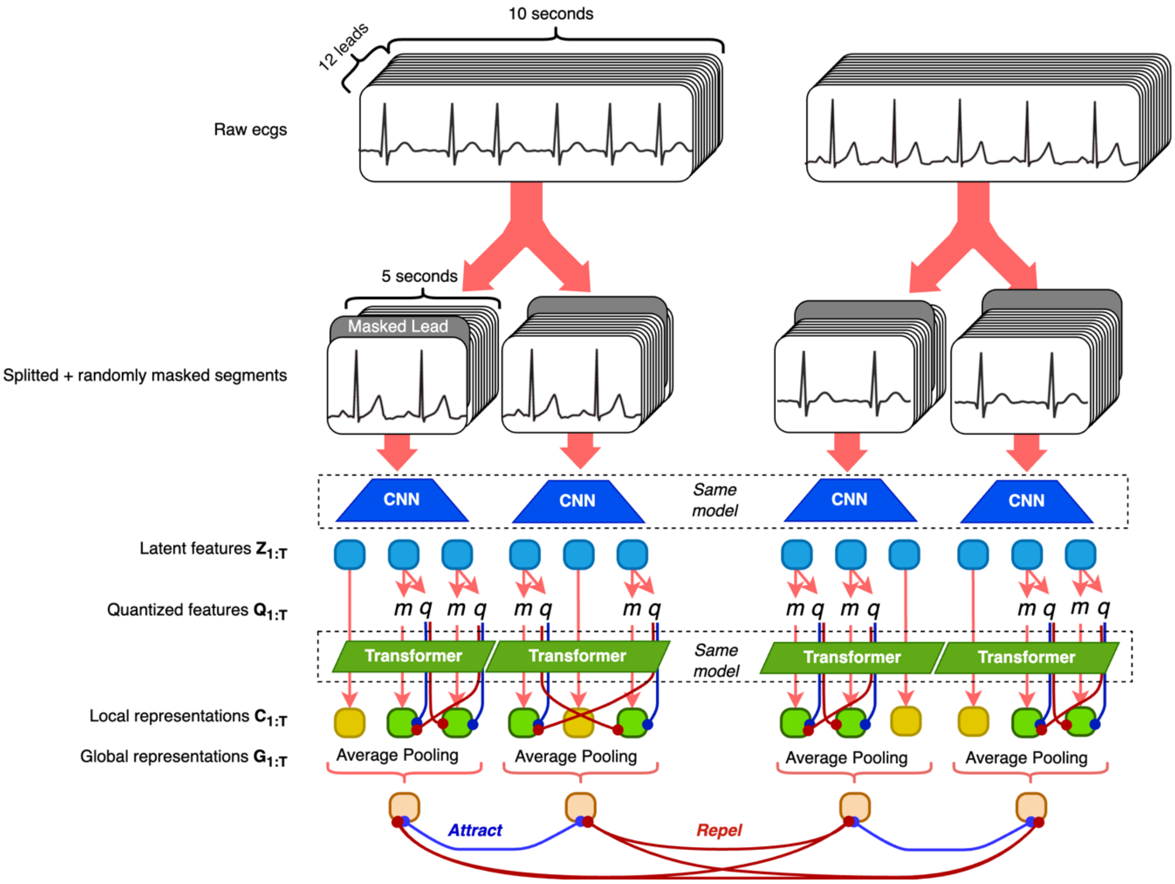


**Supplementary Figure 2: Pre-Training of DeepECG-SSL**. Raw ECGs are divided into segments, and some leads are randomly masked (zeroed out). These segments are then processed by a convolutional encoder. Within the encoder, randomly selected features are replaced with a masked token m as well as quantized into **q**. Both the masked and unmasked tokens are passed through a transformer encoder, which learns contextual representations **c**. Two contrastive losses guide this process, illustrated by blue (attraction) and red (repulsion) lines. **The intra-segment contrastive loss** encourages the contextualized representation of a token to align closely with its corresponding quantized representation within the same segment. **The inter-segment contrastive loss** operates on global representations obtained by averaging contextual representations. It pulls together global representations originating from the same ECG or the same patient, while pushing apart global representations from other ECGs within the batch. The figure is inspired by Oh et al.[42]

***Abbreviations***: **ECG:** Electrocardiogram

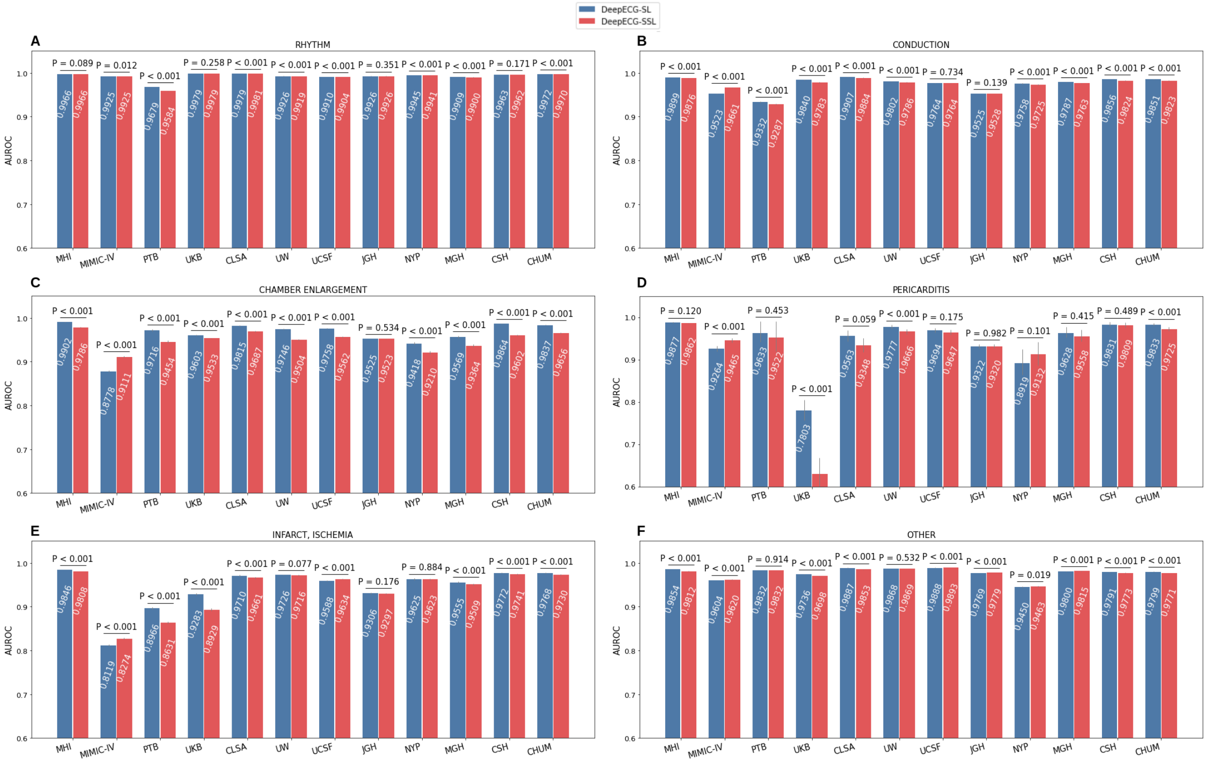


**Supplementary Figure 3: An overview of DeepECG-SL and DeepECG-SSL performances on ECG interpretation categories.** ***Abbreviations***: **ECG:** Electrocardiogram, **MHI**: Montreal Heart Institute, **CLSA:** Canadian Longitudinal Study on Aging , **UKB:** UK Biobank, **PTB**: Physikalisch-Technische Bundesanstalt, **MIMIC-IV**: Medical Information Mart for Intensive Care IV, **EPD**: External public dataset (CLSA, PTB, UKB, MIMIC-IV), **UCSF:** University of California San Francisco Medical Center**, UW:** University of Washington Medical Center**, NYP:** New York-Presbyterian Hospital, **JGH:** Jewish General Hospital**, MGH:** Massachusetts General Hospital**, CSH:** Cedars Sinai Hospital. **, EHC:** external private health center datasets (UCSF, UW, NYP, JGH, MGH, CSH)

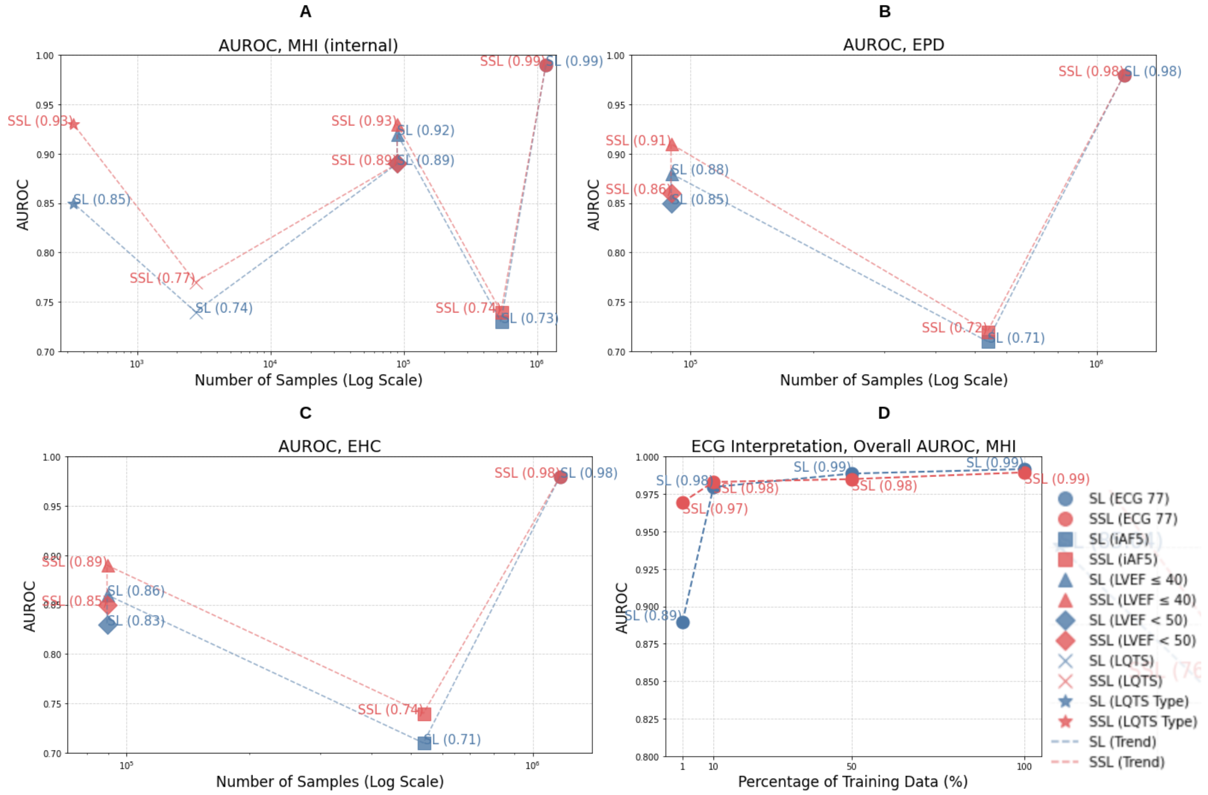


**Supplementary Figure 4: AUROC of DeepECG-SL and DeepECG-SSL as a function of the training dataset size.** ECG 77 corresponds to the ECG interpretation task. $N_{ECG77}$ = 1,166,896, $N_{LVEF}$= 537,742, $N_{iAF5}$ = 89,500, $N_{LQTS}$ = 2741, $N_{LQTStype}$ = 334. **A** AUROC on internal dataset MHI. **B** AUROC on external public dataset EPD. **C** AUROC on external private health center dataset EHC. ***Abbreviations***: **AUROC:** Area Under the Receiver Operating Characteristics Curve, **ECG**: Electrocardiogram, **MHI**: Montreal Heart Institute, **EHC:** external private health center datasets (UCSF, UW, NYP, JGH, MGH, CSH), $N_{ECG77}$: Number of ECG in the ECG interpretation dataset, $N_{iAF5}$: Number of ECG in the incidental 5 year atrial fibrillation prediction dataset. $N_{LQTS}$ Number of ECG in the LQTS detection dataset, $N_{LQTSType}$ Number of ECG in the LQTS type classification dataset, **SSL:** Self-supervised learning**, SL:** Supervised Learning. **D**. Analysis of the model's performance when trained or fine-tuned with varying percentages of the original MHI-ds-train dataset. Performance is assessed on the ECG interpretation task.

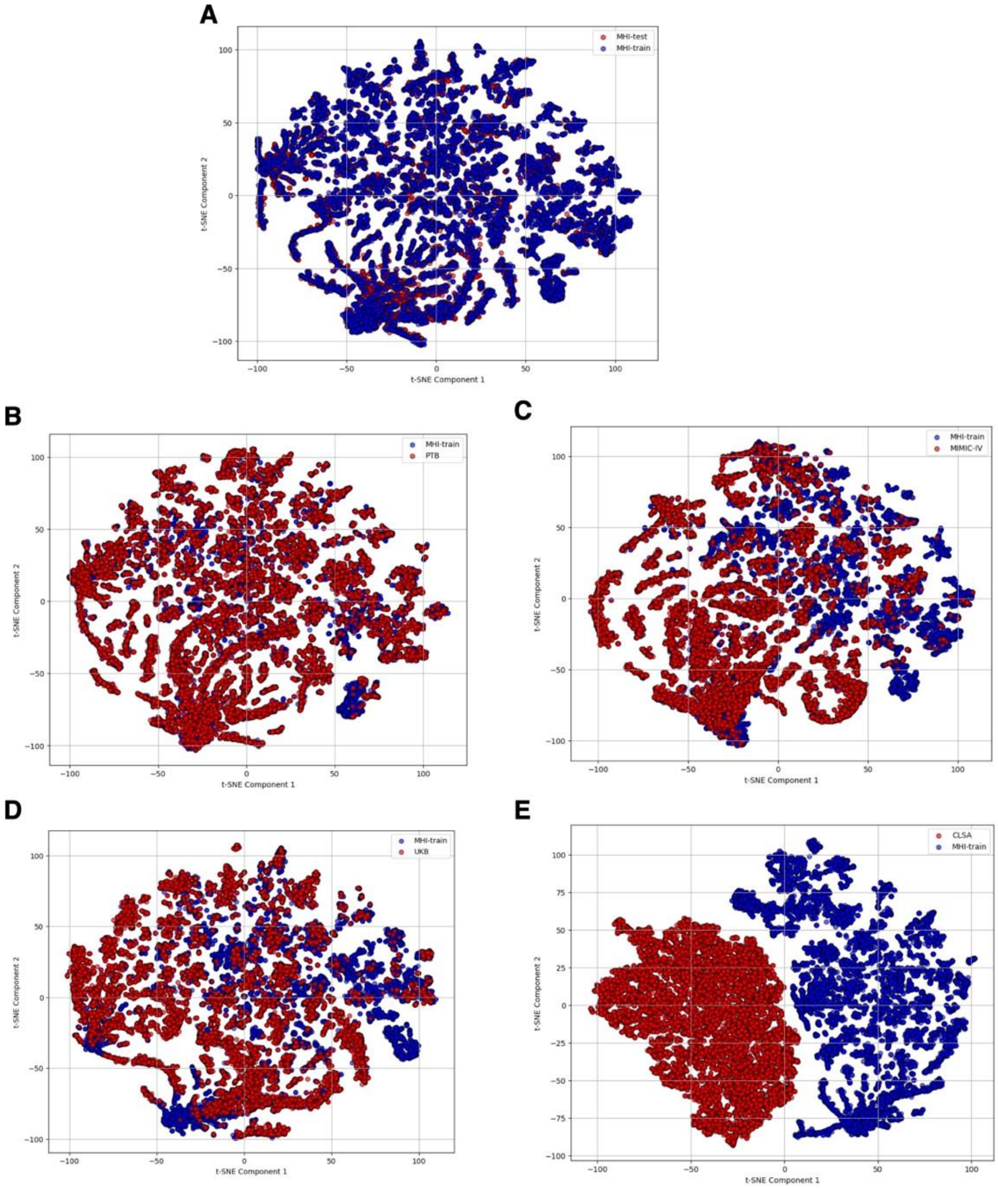


**Supplementary Figure 5 t-SNE of logits comparing each membership inference attack task for DeepECG-SL.** **A** MHI-ds-train vs MHI-ds-test, **B** MHI-ds-train vs PTB, **C** MHI-ds-train vs MIMIC-IV, **D** MHI-ds-train vs UKB, **E** MHI-ds-train vs CLSA

***Abbreviations:*** **t-SNE**: t-Distributed Stochastic Neighbor Embedding, **MHI-ds**: Montreal Heart Institute Dataset **MHI-ds-train**: MHI-ds split for training **MHI-ds-test**: **MHI-ds** split for testing **PTB**: Physikalisch-Technische Bundesanstalt, **MIMIC-IV**: Medical Information Mart for Intensive Care IV, **UKB**: UK Biobank, **CLSA**: Canadian Longitudinal Study on Aging.

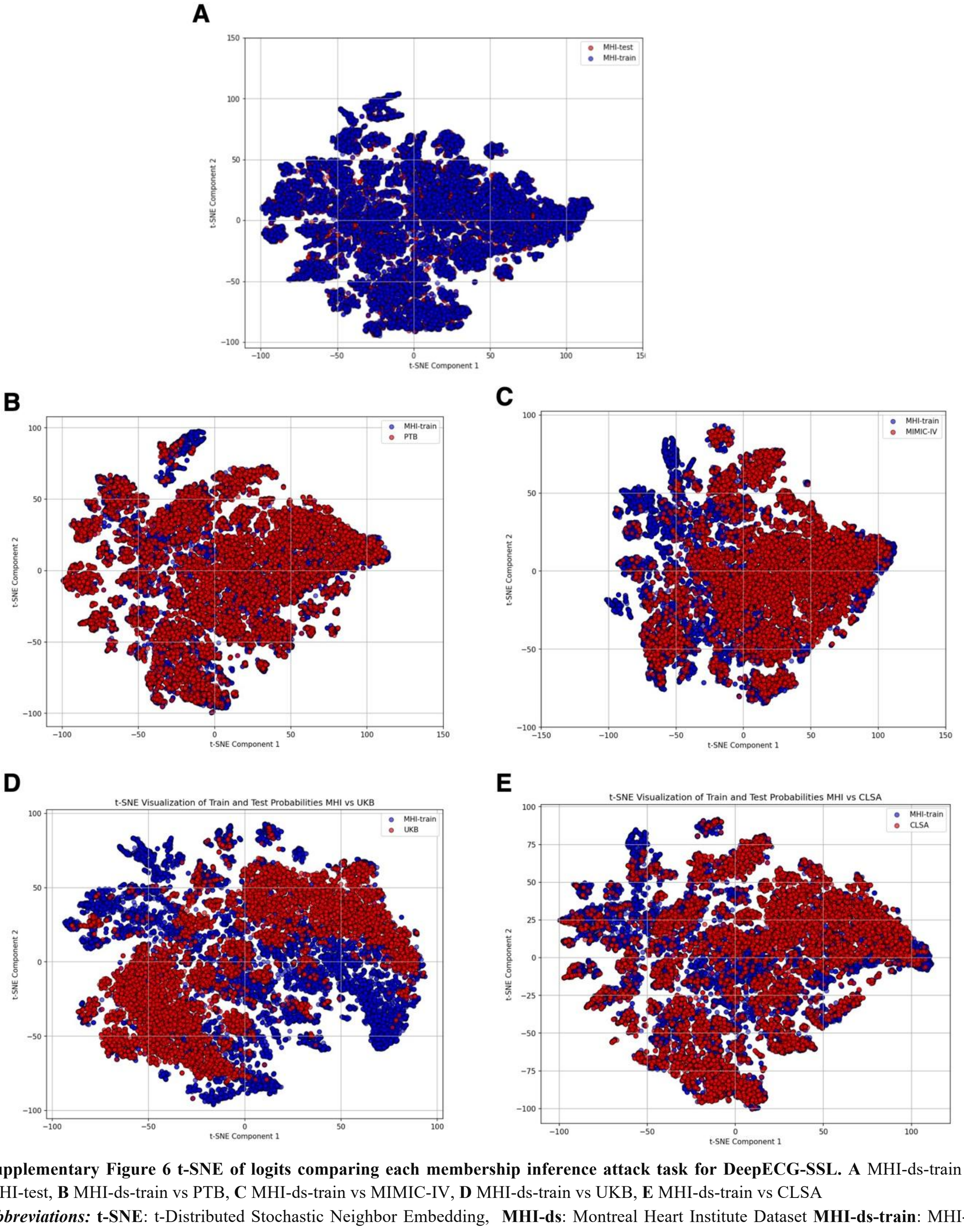


**Supplementary Figure 6 t-SNE of logits comparing each membership inference attack task for DeepECG-SSL. A** MHI-ds-train vs MHI-test, **B** MHI-ds-train vs PTB, **C** MHI-ds-train vs MIMIC-IV, **D** MHI-ds-train vs UKB, **E** MHI-ds-train vs CLSA

***Abbreviations:*** **t-SNE**: t-Distributed Stochastic Neighbor Embedding, **MHI-ds**: Montreal Heart Institute Dataset **MHI-ds-train**: MHI-ds split for training **MHI-ds-test**: **MHI-ds** split for testing **PTB**: Physikalisch-Technische Bundesanstalt, **MIMIC-IV**: Medical Information Mart for Intensive Care IV, **UKB**: UK Biobank, **CLSA**: Canadian Longitudinal Study on Aging.

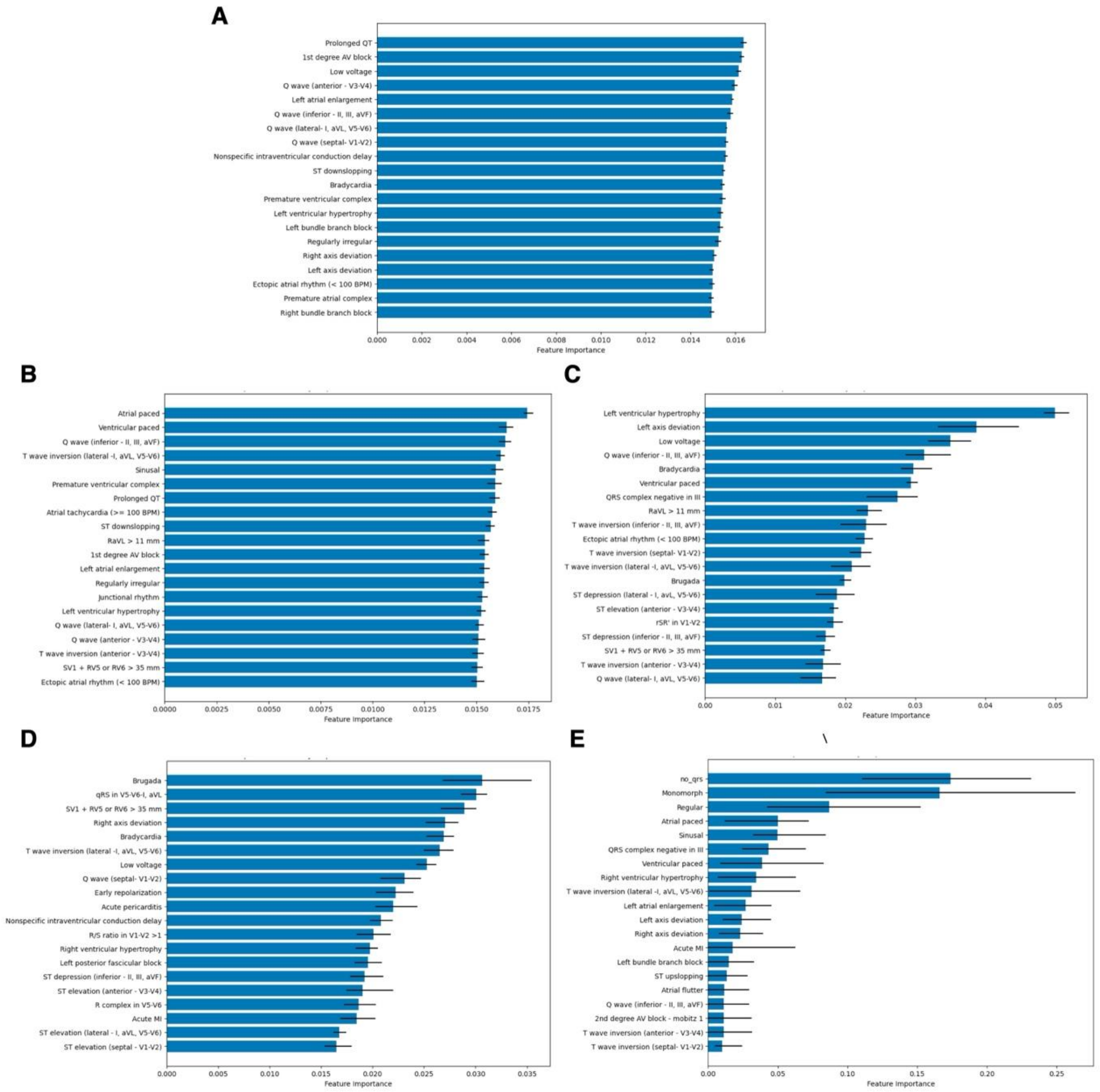


**Supplementary Figure 7 : Top 20 average feature importance for distinguishing test sets versus MHI-train for DeepECG-SL for the membership inference attack task.** Each feature is represented with 95% CI across 10 iterations of the membership inference attack against DeepECG-SL-derived logits across the datasets of interest. **A** MHI-ds-train vs MHI-ds-test, **B** MHI-ds-train vs PTB, **C** MHI-ds-train vs MIMIC-IV, **D** MHI-ds-train vs UKB, **E** MHI-ds-train vs CLSA

***Abbreviations:*** **MHI-ds**: Montreal Heart Institute Dataset **MHI-ds-train**: MHI-ds split for training **MHI-ds-test**: **MHI-ds** split for testing **PTB**: Physikalisch-Technische Bundesanstalt, **MIMIC-IV**: Medical Information Mart for Intensive Care IV, **UKB**: UK Biobank, **CLSA**: Canadian Longitudinal Study on Aging.

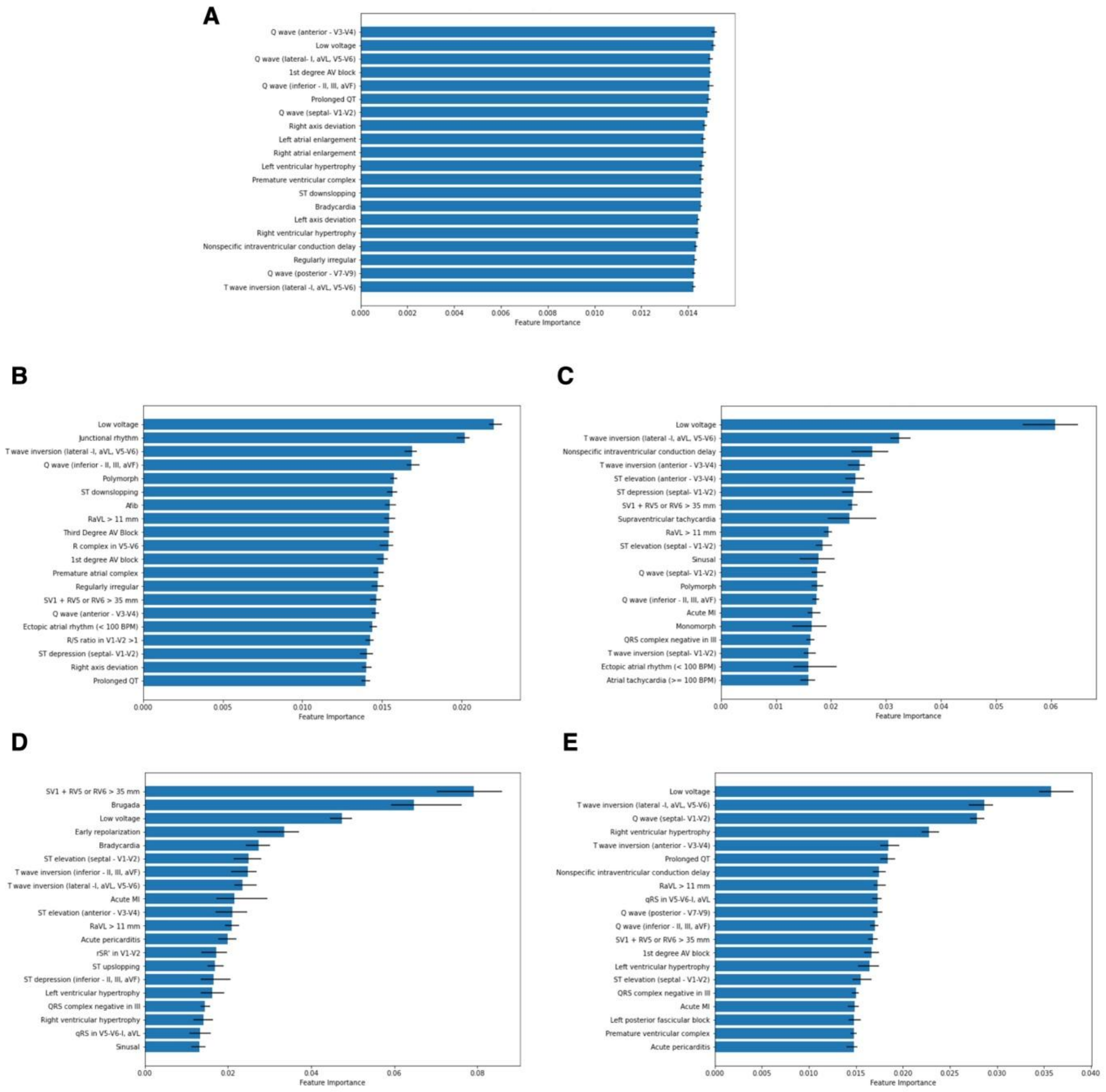


**Supplementary Figure 8 : Top 20 average feature importance for distinguishing test sets versus MHI-train for DeepECG-SSL for the membership inference attack task.** Each feature is represented with 95% CI across 10 iterations of the membership inference attack against DeepECG-SL-derived logits across the datasets of interest. **A** MHI-ds-train vs MHI-ds-test, **B** MHI-ds-train vs PTB, **C** MHI-ds-train vs MIMIC-IV,

***Abbreviations:*** **MHI-ds**: Montreal Heart Institute Dataset **MHI-ds-train**: MHI-ds split for training **MHI-ds-test**: **MHI-ds** split for testing **PTB**: Physikalisch-Technische Bundesanstalt, **MIMIC-IV**: Medical Information Mart for Intensive Care IV, **UKB**: UK Biobank, **CLSA**: Canadian Longitudinal Study on Aging.

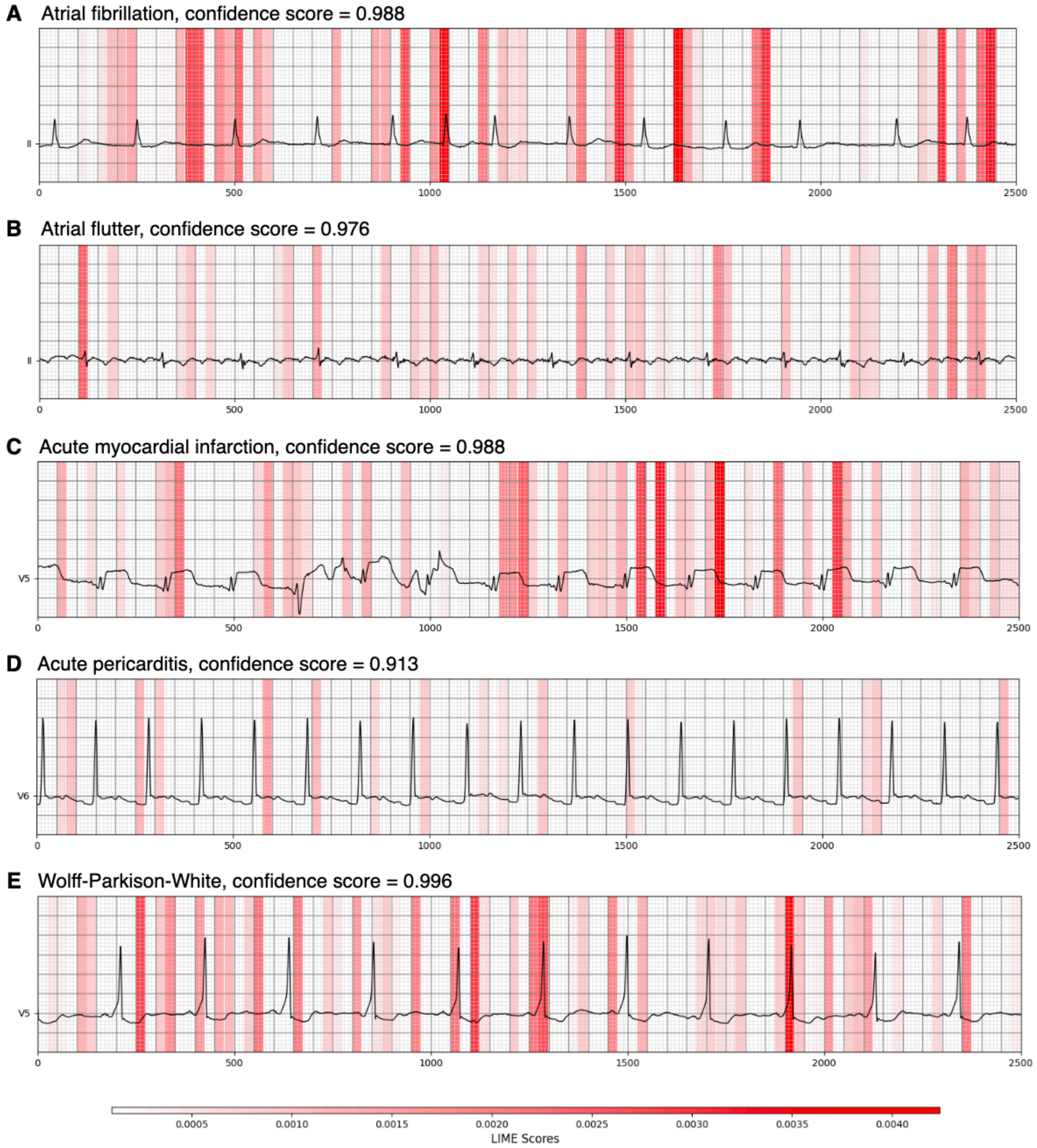


**Supplementary Figure 9: Selected examples analyzed using Local Interpretable Model-Agnostic Explanation (LIME) on DeepECG-SL only displaying positive values, highlighting the regions of greater importance for predicting the label on a chosen lead. The LIME values solely indicate how important that segment is for that particular label among the 77 labels. The confidence score represents the model's output logit's for that label following the application of the sigmoid function A.** Represents an example of atrial fibrillation. **B**. Represents an example of an atrial flutter example. **C**. Represents an acute myocardial infarction example. **D**. Represents an acute pericarditis example. **E**. Represents a Wolff-Parkinson-White example.

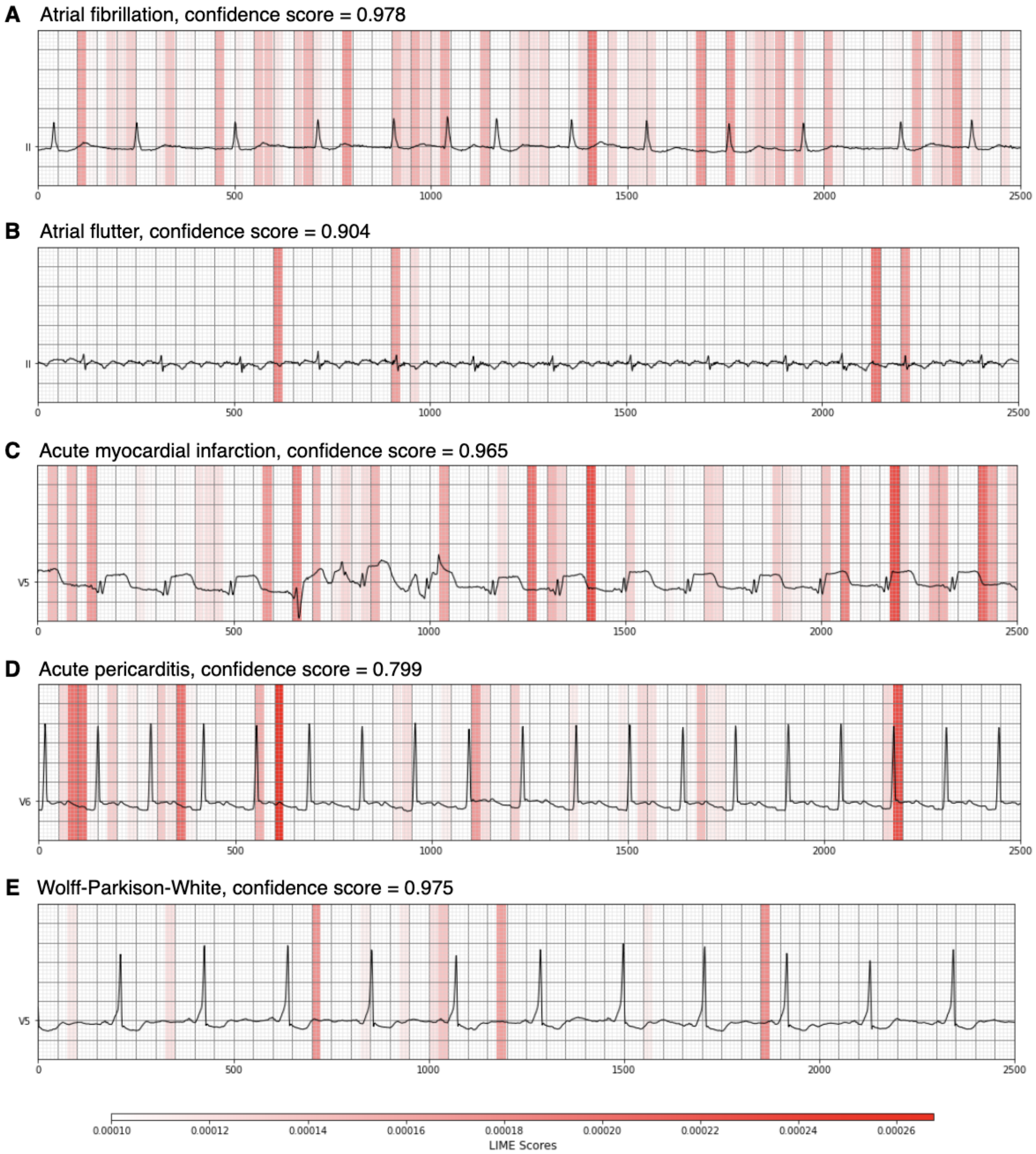


**Supplementary Figure 10: Selected examples analyzed using Local Interpretable Model-Agnostic Explanation (LIME) on DeepECG-SSL only displaying positive values, highlighting the regions of greater importance for predicting the label on a chosen lead. The LIME values solely indicate how important that segment is for that particular label among the 77 labels. The confidence score represents the model's output logit's for that label following the application of the sigmoid function A.** Represents an example of atrial fibrillation. **B**. Represents an example of an atrial flutter example. **C**. Represents an acute myocardial infarction example. **D**. Represents an acute pericarditis example. **E**. Represents a Wolff-Parkinson-White example.

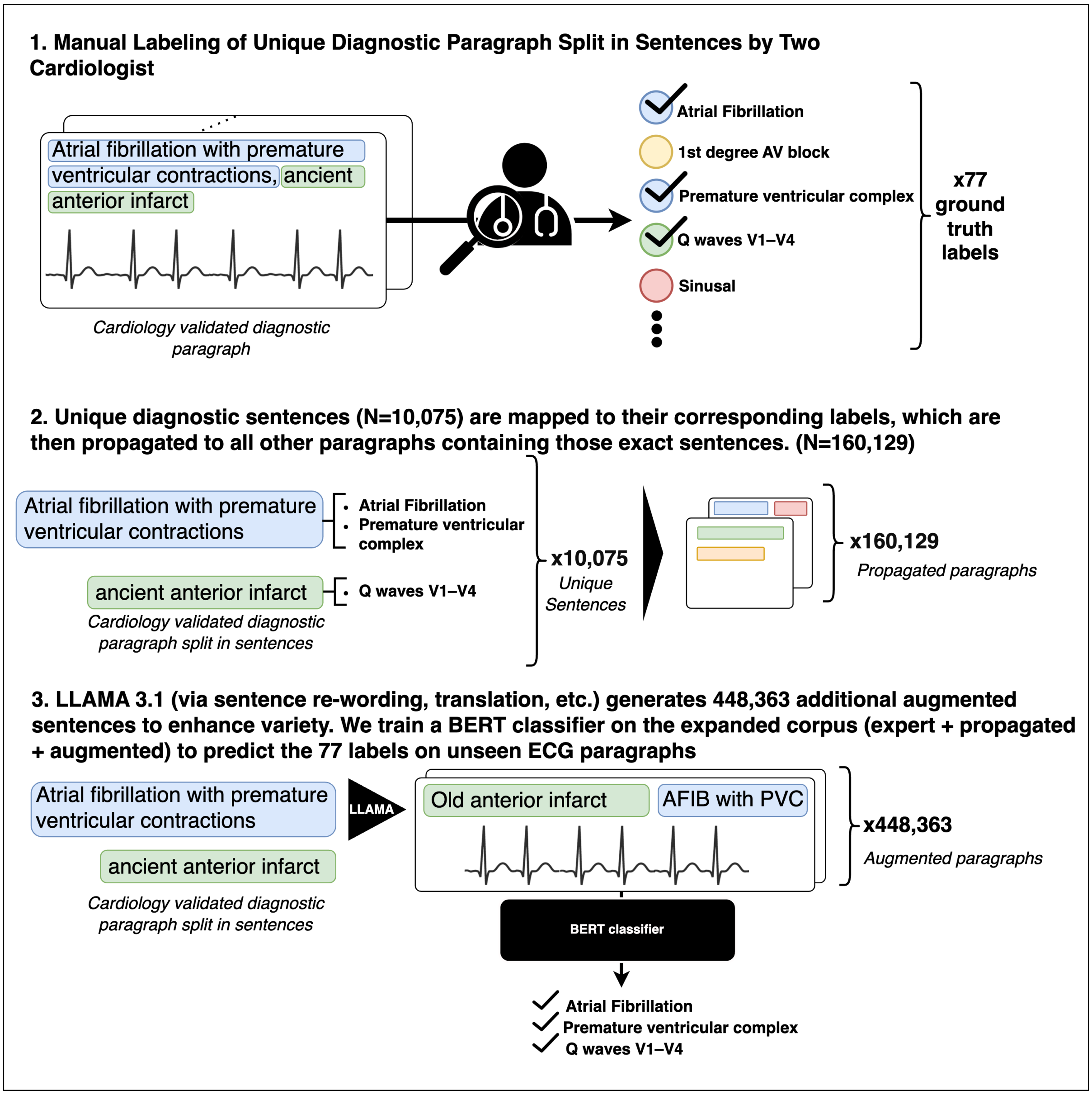


**Supplementary Figure 11: Detailed process used for the annotation of the ECGs across our datasets.**

**Supplementary Table 2: 77 labels and categories used as target labels for the ECG interpretation task.** These are grouped into six categories: Rhythm, Conduction, Chamber Enlargement, Pericarditis, Infarct-Ischemia, and Other.

| Category | Labels |
|---|---|
| RHYTHM | Ventricular tachycardia, Bradycardia, Brugada, Wolff-ParkinsonWhite (Pre-excitation syndrome), Atrial flutter, Ectopic atrial rhythm (< 100 BPM), Atrial tachycardia (>100 BPM), Sinusal, Ventricular Rhythm, Supraventricular tachycardia, Junctional rhythm, Regular, Regularly irregular, Irregularly irregular, Afib, Premature ventricular complex, Premature atrial complex |
| CONDUCTION | Left anterior fascicular block, Delta wave, 2nd degree AV block mobitz 2, Left bundle branch block, Right bundle branch block, Left axis deviation, Atrial paced, Right axis deviation, Left posterior fascicular block, 1st degree AV block, Right superior axis, Nonspecific intraventricular conduction delay, Third Degree AV Block, 2nd degree AV block - mobitz 1, Prolonged QT, U wave, LV pacing, Ventricular paced |
| CHAMBER ENLARGEMENT | Bi-atrial enlargement, Left atrial enlargement, Right atrial enlargement, Left ventricular hypertrophy, Right ventricular hypertrophy |
| PERICARDITIS | Acute pericarditis |
| INFARCT, ISCHEMIA | Q wave (septal- VI-V2), ST elevation (anterior - V3-V4), Q wave (posterior - V7-V9), Q wave (inferior - 11, 111, aVF), Q wave (anterior - V3-V4), ST elevation (lateral - 1, aVL, V5-V6), Q wave (lateral- 1, aVL, V5-V6), ST depression (lateral - 1, aVL, V5-V6), Acute MI, ST elevation (septal - VI-V2), ST elevation (inferior 11, 111, aVF), ST elevation (posterior - V7-V8-V9), ST depression (inferior - 11, 111, aVF), ST depression (anterior - V3-V4) |
| OTHER | ST downsloping, ST depression (septal- VI-V2), R/S ratio in VlV2 > 1, RVI + SV6 > 11 mm, Polymorph, rSR in VI-V2, QRS complex negative in 111, (IRS in V5-V6-1, aVL, QS complex in VlV2-V3, R complex in V5-V6, RaVL > 11 mm, T wave inversion (septal- VI-V2), SVI + RV5 or RV6 > 35 mm, T wave inversion (inferior - 11, 111, aVF), Monomorph, T wave inversion (anterior V3-V4), T wave inversion (lateral - 1, aVL, V5-V6), Low voltage, Lead misplacement, ST depression (anterior - V3-V4), Early repolarization, ST upsloping, no-qrs |

***Abbreviations***: **BPM**: beats per minute

**Supplementary Table 3: BERT Classification performance across ECG interpretation labels and categories:** These results were obtained by evaluating the model on 20% of a merged dataset comprising MIMIC-IV, MHI-ds, and PTB..

| Labels | AUROC | AUPRC | F1 Score |
|---|---|---|---|
| Ventricular tachycardia | 1.00 | 0.995 | 0.86 |
| Bradycardia | 1.00 | 1.00 | 1.00 |
| Brugada | 0.999 | 0.678 | 0.079 |
| Wolff-Parkinson-White (Pre-excitation syndrome) | 1.00 | 0.992 | 0.984 |
| Atrial flutter | 1.00 | 0.997 | 0.982 |
| Ectopic atrial rhythm (< 100 BPM) | 1.00 | 0.999 | 0.902 |
| Atrial tachycardia (>100 BPM) | 1.00 | 0.999 | 0.987 |
| Sinusal | 1.00 | 1.00 | 0.999 |
| Ventricular Rhythm | 1.00 | 0.999 | 0.992 |
| Supraventricular tachycardia | 1.00 | 0.982 | 0.52 |
| Junctional rhythm | 1.00 | 0.986 | 0.936 |
| Regular | 0.999 | 1.00 | 0.998 |
| Regularly irregular | 1.00 | 0.999 | 0.99 |

| Labels | AUROC | AUPRC | F1 Score |
|---|---|---|---|
| Ventricular tachycardia | 1.00 | 0.995 | 0.86 |
| Bradycardia | 1.00 | 1.00 | 1.00 |
| Brugada | 0.999 | 0.678 | 0.079 |
| Irregularly irregular | 1.00 | 0.999 | 0.996 |
| Afib | 1.00 | 0.998 | 0.998 |
| Premature ventricular complex | 1.00 | 0.999 | 0.993 |
| Premature atrial complex | 1.00 | 0.999 | 0.994 |
| **RHYTHM** | **1.00** | **1.00** | **0.999** |
| Left anterior fascicular block | 1.00 | 0.999 | 0.991 |
| Delta wave | 1.00 | 0.992 | 0.969 |
| 2nd degree AV block - mobitz 2 | 1.00 | 0.998 | 0.976 |
| Left bundle branch block | 1.00 | 0.999 | 0.992 |
| Right bundle branch block | 1.00 | 1.00 | 0.997 |
| Left axis deviation | 1.00 | 0.999 | 0.996 |
| Atrial paced | 1.00 | 0.995 | 0.979 |
| Right axis deviation | 1.00 | 0.997 | 0.983 |
| Left posterior fascicular block | 1.00 | 0.999 | 0.964 |
| 1st degree AV block | 1.00 | 0.999 | 0.993 |
| Right superior axis | 1.00 | 0.923 | 0.774 |
| Nonspecific intraventricular conduction delay | 1.00 | 0.999 | 0.977 |
| Third Degree AV Block | 99.9 | 0.927 | 0.181 |
| 2nd degree AV block - mobitz 1 | 1.00 | 0.991 | 0.795 |
| Prolonged QT | 1.00 | 0.999 | 0.992 |
| U wave | 98.2 | 0.952 | 0.961 |
| LV pacing | 1.00 | 0.991 | 0.269 |
| Ventricular paced | 1.00 | 0.999 | 0.986 |
| **CONDUCTION** | **1.00** | **0.999** | **0.999** |
| Bi-atrial enlargement | 1.00 | 0.996 | 0.75 |
| Left atrial enlargement | 1.00 | 0.999 | 0.992 |
| Right atrial enlargement | 1.00 | 0.991 | 0.905 |
| Left ventricular hypertrophy | 1.00 | 1.00 | 0.999 |
| Right ventricular hypertrophy | 1.00 | 0.994 | 0.967 |
| **CHAMBER ENLARGMENT** | **1.00** | **0.999** | **0.999** |
| Acute pericarditis | 1.00 | 0.997 | 0.966 |
| PERICARDITIS | 1.00 | 0.997 | 0.999 |
| Q wave (septal- VI-V2) | 1.00 | 0.999 | 0.993 |
| ST elevation (anterior ,- V3-V4) | 1.00 | 0.99 | 0.786 |
| Q wave (posterior - V7-V9) | 1.00 | 0.994 | 0.991 |
| Q wave (inferior - 11, 111, aVF) | 1.00 | 0.998 | 0.994 |
| Q wave (anterior - V3-V4) | 1.00 | 0.999 | 0.997 |
| ST elevation (lateral - 1, aVL, V5-V6) | 1.00 | 0.997 | 0.887 |

| Labels | AUROC | AUPRC | F1 Score |
|---|---|---|---|
| Ventricular tachycardia | 1.00 | 0.995 | 0.86 |
| Bradycardia | 1.00 | 1.00 | 1.00 |
| Brugada | 0.999 | 0.678 | 0.079 |
| Q wave (lateral- 1, aVL, V5-V6) | 1.00 | 0.998 | 0.979 |
| ST depression (lateral - 1, avL, V5-V6) | 1.00 | 0.999 | 0.980 |
| Acute MI | 1.00 | 0.995 | 0.851 |
| ST elevation (septal - VI-V2) | 1.00 | 0.979 | 0.781 |
| ST elevation (inferior - 11, 111, aVF) | 1.00 | 0.985 | 0.841 |
| ST elevation (posterior - V7-V8-V9) | 1.00 | 0.935 | 0.061 |
| ST depression (inferior - 11, 111, aVF) | 1.00 | 0.996 | 0.809 |
| ST depression (anterior ,- V3-V4) | 1.00 | 0.996 | 0.992 |
| **INFARCT, ISCHEMIA** | **1.00** | **0.997** | **0.998** |
| ST downslopping | 1.00 | 0.999 | 0.995 |
| ST depression (septal- VI-V2) | 1.00 | 0.995 | 0.869 |
| R/S ratio in VI-V2 >1 | 1.00 | 0.995 | 0.987 |
| RVI + SV6 > 11 mm | 1.00 | 0.996 | 0.946 |
| Polymorph | 1.00 | 0.993 | 0.967 |
| rSR' in VI-V2 | 1.00 | 0.998 | 0.992 |
| QRS complex negative in 111 | 99.9 | 0.997 | 0.976 |
| qRS in V5-V6-1, aVL | 1.00 | 0.998 | 0.983 |
| QS complex in VI-V2-V3 | 1.00 | 0.998 | 0.996 |
| R complex in V5-V6 | 1.00 | 0.998 | 0.994 |
| RaVL > 11 mm | 1.00 | 0.999 | 0.991 |
| T wave inversion (septal- VI-V2) | 1.00 | 0.997 | 0.987 |
| SVI + RV5 or RV6 > 35 mm | 1.00 | 0.999 | 0.992 |
| T wave inversion (inferior - 11, 111, aVF) | 1.00 | 0.998 | 0.992 |
| Monomorph | 99.9 | 1.00 | 0.997 |
| T wave inversion (anterior ,- V3-V4) | 1.00 | 0.998 | 0.975 |
| T wave inversion (lateral -1, aVL, V5-V6) | 99.9 | 0.997 | 0.988 |
| Low voltage | 1.00 | 1.00 | 0.996 |
| Lead misplacement | 99.9 | 0.947 | 0.022 |
| ST depression (anterior - V3-V4) | 1.00 | 0.996 | 0.992 |
| Early repolarization | 1.00 | 0.998 | 0.988 |
| ST upslopping | 99.9 | 0.995 | 0.943 |
| no-qrs | 1.00 | 1.00 | 1.00 |
| **OTHER** | **1.00** | **0.999** | **0.998** |

***Abbreviations***: **BPM**: beats per minute, **AUROC**: Area Under the Receiver Operating Characteristics Curve, **AUPRC:** Area Under the Precision Recall Curve

**Supplementary Table 4: Equalized Odds for Gender and Age Across Categories.** The true positive rate (TPR) and false positive rate (FPR) were calculated to evaluate model performance across different subgroups. TPR measures the proportion of actual positives correctly identified, while FPR quantifies the proportion of negatives incorrectly classified as positives. Gender-based comparisons assessed performance differences between female and male patients, while age-based comparisons analyzed results for patients in three age groups: under 55, 55 to 75, and over 75 years.

| Dataset | Category | TPR | | | FPR | | |
|---|---|---|---|---|---|---|---|
| | | DeepECG-SL | DeepECG-SSL | $\Delta_{SSL-SL}$ | DeepECG-SL | DeepECG-SSL | $\Delta_{SSL-SL}$ |
| MHI-test | Gender | 0.021 [0.019,0.023] | 0.02 [0.019,0.022] | - | 0.002 [0.002,0.002] | 0.002 [0.002,0.002] | - |
| | Age | 0.04 [0.038,0.042] | 0.034 [0.033,0.036] | -0.01 | 0.005 [0.005, 0.005] | 0.005 [0.005, 0.005] | - |
| MIMIC-IV-test | Gender | 0.047 [0.040,0.054] | 0.013 [0.008,0.016] | -0.03 | 0.001 [0.001,0.002] | 0.0 [0.000,0.000] | 0 |
| | Age | 0.045 [0.038,0.050] | 0.021 [0.016,0.024] | -0.02 | 0.006 [0.006,0.006] | 0 [0.000,0.000] | -0.01 |
| PTB | Gender | 0.03 [0.026,0.034] | 0.016 [0.012,0.019] | -0.01 | 0.003 [0.003,0.004] | 0.001 [0.001,0.001] | 0 |
| | Age | 0.049 [0.042,0.060] | 0.03 [0.024,0.038] | -0.02 | 0.01 [0.009,0.010] | 0.002 [0.001,0.002] | -0.01 |
| UKB | Gender | 0.032 [0.028,0.037] | 0.018 [0.014,0.023] | -0.01 | 0.008 [0.007,0.008] | 0.0 [0.000,0.001] | -0.01 |
| | Age | 0.046 [0.040, 0.052] | 0.029 [0.025,0.034] | -0.02 | 0.009 [0.008,0.009] | 0.001 [0.001,0.0011] | -0.01 |
| CLSA | Gender | 0.04 [0.033, 0.048] | 0.017 [0.013,0.023] | -0.02 | 0.004 [0.004, 0.004] | 0.001 [0.000, 0.001] | 0 |
| | Age | 0.071 [0.064, 0.079] | 0.032 [0.026, 0.039] | -0.04 | 0.011 [0.010, 0.011] | 0.001 [0.001, 0.001] | -0.01 |

***Abbreviation***: **TPR**: True Positive Rate, **FPR**: False Positive Rate. $\Delta_{SSL-SL}$: Difference in performance between DeepECG-SSL and DeepECG-SL.

**Supplemental Table 5: Membership Inference Attack Evaluation Metrics for MIMIC-IV, UKB, MHI-ds, and PTB Datasets.** Results were obtained using an attack model trained on each comparison with MHI-train.

| Dataset | AUROC | | | AUPRC | | |
|---|---|---|---|---|---|---|
| | DeepECG-SL | DeepECG-SSL | $\Delta_{SSL-SL}$ | DeepECG-SL | DeepECG-SSL | $\Delta_{SSL-SL}$ |
| MH1-test | 57.84 [57.59,58.18] | 57.41 [57.26,57.68] | - | 55.67 [55.45,56.00] | 55.32 [55.10,55.57] | - |
| MIMIC-1V-test | 97.57 [97.51,97.60] | 95.64 [95.55,95.75] | -1.93 | 97.16 [97.10, 97.23] | 95.03 [94.89, 95.17] | -2.13 |
| PTB | 68.3 [67.80, 68.84] | 77.26 [76.83,77.85] | 8.96 | 63.1 [62.69, 63.51] | 71.0 [70.45, 71.65] | 7.9 |
| UKB | 97.81 [97.74, 97.89] | 99.08 [99.00,99.13] | 1.27 | 96.89 [96.76, 97.02] | 98.65 [98.50, 98.73] | 1.76 |
| CLSA | 100.0 [100.00,100.0] | 93.53 [93.33,93.73] | -6.47 | 100.0 [100.00,100.00] | 91.48 [91.15, 91.81] | -8.52 |

***Abbreviations***: **MIMIC-IV**:Medical Information Mart for Intensive Care Dataset IV, **UKB**: UK Biobank, **MHI-ds**: Montreal Heart Institute dataset, **PTB**: Physikalisch-Technische Bundesanstalt, **AUROC**: Area Under the Receiver Operating Characteristics Curve, **AUPRC:** Area Under the Precision Recall Curve. **CLSA:** Canadian Longitudinal Study on Aging.

**Supplementary Table 6: Comparison of DeepECG-SSL and DeepECG-SL Inference and Training Parameters.** Inference times and emissions are based on processing 1000 examples 1000 times.

| Metric | Device | DeepECG-SL | DeepECG-SSL | Order of Difference |
|---|---|---|---|---|
| Inference Time (seconds) | GPU | 0.4767 [0.4641,0.4893] | 3.1488 [1.9037, 4.3939] | 6.6x |
| | CPU | 10.2990 [10.2297, 10.3683] | 303.3232 [301.3252, 305.3212] | 29x |
| C02 Emissions (mgC02) | GPU | 0.4246 [0.4077, 0.4415] | 1.7741 [1.1859, 2.3622] | 4.2x |
| | CPU | 8.8042 [8.7281, 8.8803] | 85.4187 [81.6596, 89.1778] | 9.7x |
| Energy Consumption (Wh) | GPU | 0.1786 [0.1715, 0.1857] | 0.7463 [0.4989, 0.9937] | 4.2x |

| | | | | |
|---|---|---|---|---|
| | CPU | 3.7036<br>[3.6716, 3.7356] | 35.9322<br>[34.3509, 37.5135] | 9.7x |
| Params | - | 1.51 M | 90.37 M | 60x |
| fwd MACs | | 530.57 MMACs | 14.17 GMACs | 27x |
| fwd FLOPs | | 1.09 GFLOPS | 28.37 GFLOPS | 26x |
| fwd+bwd MACs | | 1.59 GMACs | 42.52 GMACs | 27x |
| fwd+bwd FLOPs | | 3.27 GFLOPS | 85.10 GFLOPS | 26x |

***Abbreviations***: **M**: million, **G**: billion, **MACs** : Multiply-Accumulate Operations, **FLOPs**: Floating Point Operations, **Wh**: watt-hours, **fwd**: forward pass, **fwd+bwd**: forward + backwards pass.

**Supplementary Table 7: Hyperparameters & Training environment parameters for the DeepECG-SL.**

| Parameter | Value |
|---|---|
| Activation function | leaky-relu[25] |
| Augmentation Function | magnitude-warp-uniform-multithreaded[19] |
| Architecture | s_v2[11] |
| Apply Label Smoothing[47] | True |
| Base Channels | [12, 12, 24, 32, 64, 80, 128, 640] |
| Base Depths | [1,1,2,2,3,4,5] |
| Batch Size | 780 |
| Data | unscaled |
| Dropout[31] | 0.1193 |
| EMA Value | 0.8183 |
| Expansion Factors | [1, 2, 2, 2, 2, 2, 2] |
| Kernel Sizes | [3, 3, 5, 3, 5, 3, 3, 3] |
| Label Smoothing Factor | 8.98E-08 |
| Loss | MultiLabe1SoftMarginLoss |
| LR | 0.0077 |
| Augmentation Type | Per batch |
| Max Epochs | 25 |
| Output Activation | sigmoid |
| Optimiser | AdamW[4] |
| Output Neurons | 77 |
| Rand Augment[48] | 0.1849 |
| Scheduler | cosine_annealing[37] |
| SE Ratio | [4, 4, 4, 4, 4, 4, 4] |
| Strides | [1, 1, 2, 2, 2, 2, 2, 2] |
| Stochastic Depth[30] | 0.304 |
| Use Adaptive Clipping[38] | True |
| Use EMA | False |
| Use SE | True |
| Use Warmup | True |
| Weight Decay | 0.00031 |

***Abbreviation***: **EMA**: Exponential Moving Average, **LR**: Learning Rate, **SE**: Squeeze-and-Excitation**.** Other terms such as **AdamW**, **cosine annealing**, and **MultiLabelSoftMarginLoss** refer to specific optimizer, scheduler, and loss function types. All parameter names follow standard conventions in neural network training.

**Supplementary Table 8: Optimization Parameters for DeepECG-SL:** Each model family had its sub-variations tested. For transformers, parameters such as patch size, MLP dimension, and number of heads were varied, while convolutional networks had their kernel size, stride size, and normalization strategy optimized.

| Parameter | Options |
|---|---|
| Scaling Method | MinMax, Quantile, Standard Scale, Robust Scaling, No Scaling Per Example, Per Lead, Per Dataset |
| Scaling Granularity | Per Example, Per Lead, Per Dataset |
| Rand Augment[48] | [0.0, 0.7] |
| Augment Function | window-warp-multithreaded, window-slice multithreaded, time-warp-multithreaded, magnitude-warp-uniform multithreaded, beat-permutation, scaling, jitter, none[25] |
| Learning Rate | [0.01, 0.000000001] |
| Scheduler | by-plateau, cosine-annealing[37], none, triangular2, lambda |
| Optimiser | Adam[32], AdamW[4], Radam[33], SGD[34], Adagrad[35], and RMSprop[36] |
| Activation Function | ReLU[24], Leaky ReLU[25], GELU[26], SELU[27], Mish[28], and Swish[29] |
| Max Epochs | 50 |
| EMA Value | [0.9999999, 0.80] |
| Apply Label Smoothing[47] | True, False |
| Label Smoothing Factor[47] | [0.0000001, 0.0] |
| Rand Augment[48] | False, True |
| Stochastic Depth[30] | [0.5, 0.0] |
| Weight Decay | [0.001, 0.0000001] |
| Dropout[31] | [0.0, 0.5] |
| Batch Size | 218, 512, 780, 1024 |
| Output Neurons | 77 |
| Output Activation | sigmoid |
| Use Adaptive Clipping[38] | True, False |
| Use Warmup | True, False |
| Use EMA | True, False |
| Loss Functions | TwoWayLoss[21], Hill Loss[22], Asymmetric Loss[23], SPLC[22], MultiLabelSoftMarginLoss, Binary Cross-Entropy (binary-ce), Dice Loss, Binary Focal Loss (gamma=2), Binary Focal Loss (gamma=3)[3], and Weighted Binary Cross-Entropy (weighted-bce) |
| Model Families | ViT[8], CrossViT[9], EfficientNet[10], [11] , Mamba[12], ResNet[13], ResNeXt[13], Inception[15], DenseNet[16] |

**Supplementary Table 9: Optimization Parameters for DeepECG-SSL** Limited computational resources prevented us from exploring all possible combinations. For SIMCLR and BYOL, we tested ResNet50 and ResNet101 using the same set of data augmentations. For JEPA, we tested a 1D Transformer (1D patch + ViT base). For WCR, we applied the configuration outlined in Oh et al.[42].

| Parameter | Options |
|---|---|
| Strategies | BYOL[40] , JEPA[41], SIMCLR[39], WCR[42] |
| Data transformation | gaussian blur, gaussian noise, em noise, pl noise, random lead mask, sobel derivative, random wanderer, baseline wanderer, baseline shift, scaling, none |
| Max Epochs | 100, 200, 250 |

| | |
|---|---|
| Batch Size | 1024 |
| Model Families | ViT[8], ResNet[13] |

***Abbreviations***: **BYOL** (Bootstrap Your Own Latent), **JEPA** (Joint Embedding Predictive Architecture), **SIMCLR** (Simple Framework for Contrastive Learning of Visual Representations), **WCR** (Wave2Vec+Contrastive Multi-Segment Coding+Random Lead Masking)

**Supplementary Table 10: Performance Metrics Comparison Across Cleaned and Raw Datasets (MHI, MIMIC-IV, PTB, UKB, CLSA)**
**BPM**: Beats per minute, **Afib:** Atrial Fibrillation
**(See excel file: DeepECG_Heartwise_Manuscript_supp_tables.xlsx)**

**Supplementary Table 11 : Performance of the models on the MHI dataset (N = 287,039).** When the CI are non-overlapping, the difference is reported in ΔSSL-SL. Prevalence for label groups counts any positive in a member of its class as a positive.
***Abbreviations*** **ΔSSL-SL**: score difference between DeepECG-SSL and DeepECG-SL, **BPM**: Beats per minute, **Afib:** Atrial Fibrillation
**(See excel file: DeepECG_Heartwise_Manuscript_supp_tables.xlsx)**

**Supplementary Table 12: Performance of the models on the MIMIC-IV dataset (N = 242,349).** When the CI are non-overlapping, the difference is reported in ΔSSL-SL. Prevalence for label groups counts any positive in a member of its class as a positive.
**(See excel file: DeepECG_Heartwise_Manuscript_supp_tables.xlsx)**

**Supplementary Table 13: Performance of the models on the PTB dataset (N = 21,799).** When the CI are non-overlapping, the difference is reported in ΔSSL-SL. Prevalence for label groups counts any positive in a member of its class as a positive.
***Abbreviations*** **ΔSSL-SL**: score difference between DeepECG-SSL and DeepECG-SL, **BPM**: Beats per minute, **Afib:** Atrial Fibrillation
**(See excel file: DeepECG_Heartwise_Manuscript_supp_tables.xlsx)**

**Supplementary Table 14: Performance of the models on the UKB dataset (N = 54,959)** When the CI are non-overlapping, the difference is reported in ΔSSL-SL. Prevalence for label groups counts any positive in a member of its class as a positive.
***Abbreviations*** **ΔSSL-SL**: score difference between DeepECG-SSL and DeepECG-SL, **BPM**: Beats per minute, **Afib**: Atrial Fibrillation
**(See excel file: DeepECG_Heartwise_Manuscript_supp_tables.xlsx)**

**Supplementary Table 15: Performance of the models on the CLSA dataset (N = 54,758).**When the CI are non-overlapping, the difference is reported in ΔSSL-SL. Prevalence for label groups counts any positive in a member of its class as a positive.
***Abbreviations*** **ΔSSL-SL**: score difference between DeepECG-SSL and DeepECG-SL, **BPM**: Beats per minute, **Afib**: Atrial Fibrillation
**(See excel file: DeepECG_Heartwise_Manuscript_supp_tables.xlsx)**

**Supplementary Table 16: Performance of the models on the EPD dataset (N = 373,865).** When the CI are non-overlapping, the difference is reported in ΔSSL-SL. Prevalence for label groups counts any positive in a member of its class as a positive.
***Abbreviations*** **ΔSSL-SL**: score difference between DeepECG-SSL and DeepECG-SL, **EPD**: External public datasets
**(See excel file: DeepECG_Heartwise_Manuscript_supp_tables.xlsx)**

**Supplementary Table 17: Performance of the models on the UW dataset (N = 63,838).** When the CI are non-overlapping, the difference is reported in ΔSSL-SL. Prevalence for label groups counts any positive in a member of its class as a positive.
***Abbreviations*** **ΔSSL-SL**: score difference between DeepECG-SSL and DeepECG-SL, **UW**: University of Washington Dataset
**(See excel file: DeepECG_Heartwise_Manuscript_supp_tables.xlsx)**

**Supplementary Table 18: Performance of the models on the UCSF dataset (N = 108,479).** When the CI are non-overlapping, the difference is reported in ΔSSL-SL. Prevalence for label groups counts any positive in a member of its class as a positive.
***Abbreviations*** **UCSF**: University of California San Francisco
**(See excel file: DeepECG_Heartwise_Manuscript_supp_tables.xlsx)**

**Supplementary Table 19: Performance of the models on the JGH dataset (N = 218,776)** When the CI are non-overlapping, the difference is reported in ΔSSL-SL. Prevalence for label groups counts any positive in a member of its class as a positive.
***Abbreviations*** **JGH**: Jewish General Hospital
**(See excel file: DeepECG_Heartwise_Manuscript_supp_tables.xlsx)**

**Supplementary Table 20: Performance of the models on the NYP dataset (N = 10,000).** When the CI are non-overlapping, the difference is reported in ΔSSL-SL. Prevalence for label groups counts any positive in a member of its class as a positive.
***Abbreviations*** **NYP**: New York-Presbyterian Hospital
**(See excel file: DeepECG_Heartwise_Manuscript_supp_tables.xlsx)**

**Supplementary Table 21: Performance of the models on the MGH dataset (N = 20,000).** When the CI are non-overlapping, the difference is reported in ΔSSL-SL. Prevalence for label groups counts any positive in a member of its class as a positive.
***Abbreviations*** **MGH**: Massachusetts General Hospital
**(See excel file: DeepECG_Heartwise_Manuscript_supp_tables.xlsx)**

**Supplementary Table 22: Performance of the models on the CSH dataset (N = 26,445).** When the CI are non-overlapping, the difference is reported in ΔSSL-SL. Prevalence for label groups counts any positive in a member of its class as a positive.
***Abbreviations*** **CSH**: Cedars Sinai Hospital

**(See excel file: DeepECG_Heartwise_Manuscript_supp_tables.xlsx)**

**Supplementary Table 23: Performance of the models on the EHC dataset (N = 447,538).** When the CI are non-overlapping, the difference is reported in ΔSSL-SL. Prevalence for label groups counts any positive in a member of its class as a positive.
***Abbreviations*** **EHC**: External private health centers
**(See excel file: DeepECG_Heartwise_Manuscript_supp_tables.xlsx)**

**Table 24: Metrics for literature models are presented as reported in their respective publications for the corresponding datasets.**

| Datasets | Tasks | Source | AUROC | AUPRC | MAE | R2 |
|---|---|---|---|---|---|---|
| **MHI** | iAF5 | DeepECG-SL | 0.734 (0.731, 0.737) | 0.297 (0.292, 0.301) | - | - |
| | | DeepECG-SSL | 0.742 (0.739, 0.745) | 0.316 (0.311, 0.321) | - | - |
| | | Jabbour et al. | 0.75 (0.745, 0.753) | 0.31 (0.303, 0.318) | - | - |
| | LQTS | DeepECG-SL | 0.735 (0.704, 0.768) | 0.409 (0.346, 0.463) | - | - |
| | | DeepECG-SSL | 0.767 (0.736, 0.798) | 0.499 (0.445, 0.55) | - | - |
| | | Jiang et al. | 0.78 (0.76, 0.80) | - | - | - |
| **MIMIC-IV** | LVEF Regression | DeepECG-SL | - | - | 7.49 (7.37, 7.61) | 45.82 (44.03, 47.70) |
| | | DeepECG-SSL | - | - | 7.26 (7.16, 7.36) | 55.61 (54.33, 56.83) |
| | | Hou et al. | - | - | 9.56 (9.15, 9.98) | 37.20 (31.60, 42.10) |
| | | | | | 7.012 | |
| | LVEF < 50 | DeepECG-SL | 0.849 (0.842, 0.856) | 0.748 (0.735, 0.761) | - | - |
| | | DeepECG-SSL | 0.864 | 0.788 | - | - |

| | | | | | | |
|---|---|---|---|---|---|---|
| | | | (0.857, 0.87) | (0.777, 0.799) | | |
| | | Hou et al. | 0.848 | - | - | - |
| | | ECGFounder | 0.867<br>(0.86, 0.874) | | | |
| | iAF5 | DeepECG-SL | 0.711<br>(0.71, 0.713) | 0.311<br>(0.386, 0.313) | - | - |
| | | DeepECG-SSL | 0.715<br>(0.714, 0.716) | 0.326<br>(0.323, 0.328) | - | - |
| | | Jabbour et al. | 0.71<br>(0.704, 0.708) | 0.3<br>(0.296, 0.303) | - | - |

**Supplementary Table 25: Performance of DeepECG-SL and DeepECG-SSL for digital biomarker prediction on the MHI dataset** When the difference is significantly different, the difference is highlighted as ΔSSL-SL.
**(See excel file: DeepECG_Heartwise_Manuscript_supp_tables.xlsx)**

**Supplementary Table 26: Performance of DeepECG-SL and DeepECG-SSL for digital biomarker prediction on the MIMIC-IV dataset** When the difference is significantly different, the difference is highlighted as ΔSSL-SL.
**(See excel file: DeepECG_Heartwise_Manuscript_supp_tables.xlsx)**

**Supplementary Table 27: Performance of DeepECG-SL and DeepECG-SSL for digital biomarker prediction on the EPD dataset** When the difference is significantly different, the difference is highlighted as ΔSSL-SL.
**(See excel file: DeepECG_Heartwise_Manuscript_supp_tables.xlsx)**

**Supplementary Table 28: Performance of DeepECG-SL and DeepECG-SSL for digital biomarker prediction on the UW dataset** When the difference is significantly different, the difference is highlighted as ΔSSL-SL.
**(See excel file: DeepECG_Heartwise_Manuscript_supp_tables.xlsx)**

**Supplementary Table 29: Performance of DeepECG-SL and DeepECG-SSL for digital biomarker prediction on the UCSF dataset** When the difference is significantly different, the difference is highlighted as ΔSSL-SL.
***Abbreviations*** **UCSF**: University of California San Francisco
**(See excel file: DeepECG_Heartwise_Manuscript_supp_tables.xlsx)**

**Supplementary Table 30: Performance of DeepECG-SL and DeepECG-SSL for digital biomarker prediction on the JGH dataset** When the difference is significantly different, the difference is highlighted as ΔSSL-SL.
***Abbreviations*** **JGH**: Jewish General Hospital
**(See excel file: DeepECG_Heartwise_Manuscript_supp_tables.xlsx)**

**Supplementary Table 31: Performance of DeepECG-SL and DeepECG-SSL for digital biomarker prediction on the NYP dataset** When the difference is significantly different, the difference is highlighted as ΔSSL-SL.

***Abbreviations*** **NYP**: New York-Presbyterian Hospital

**(See excel file: DeepECG_Heartwise_Manuscript_supp_tables.xlsx)**

**Supplementary Table 32: Performance of DeepECG-SL and DeepECG-SSL for digital biomarker prediction on the CSH dataset** When the difference is significantly different, the difference is highlighted as ΔSSL-SL.
***Abbreviations*** **CSH**: Cedars Sinai Hospital

**(See excel file: DeepECG_Heartwise_Manuscript_supp_tables.xlsx)**

**Supplementary Table 33: Performance of DeepECG-SL and DeepECG-SSL for digital biomarker prediction on the EHC dataset** When the difference is significantly different, the difference is highlighted as ΔSSL-SL.
***Abbreviations*** **EHC**: External private health centers

**(See excel file: DeepECG_Heartwise_Manuscript_supp_tables.xlsx)**

**Supplemental Table 34 . Detailed Architecture Summary for DeepECG-SL**

```
=================================================================
Layer (type:depth-idx)                    Param #
=================================================================
├─Conv1d: 2-1                             432
├─CustomNorm: 2-2                         --
│    └─BatchNorm1d: 3-1                   24
├─LeakyReLU: 2-3                          --
├─FusedMBConv1d (x4): 2-4-2-9             --
│    └─Sequential: 3-2, 3-6, 3-10, 3-14   456, 456, 912, 1776
│    └─StochasticDepth: 3-3-7-15-19-23    --
│    └─SEBlock: 3-4-8-12-16-20            87, 87, 318, 318, 318, 318
│    └─Dropout: 3-5-9-13-17-21            --
=================================================================
```

***Abbreviations***: **Conv1d** for 1-dimensional convolution, **CustomNorm** for custom normalization, **BatchNorm1d** for 1-dimensional batch normalization, **LeakyReLU** for the Leaky Rectified Linear Unit activation function, **FusedMBConv1d** for fused MobileNetV1-like 1D convolution block, **Sequential** for stacking layers in sequence, **StochasticDepth** for layer dropping during training, **SEBlock** for Squeeze-and-Excitation blocks.

**Supplemental Table 35. Detailed cohort overview**

| **Dataset** | | **MHI-ds-train** | **MHI-ds-val** | **MHI-ds-test** | **MIMIC-IV-train** | **MIMIC-IV-test** | **PTB** | **UKB** | **CLSA** | **CODE-15** |
|---|---|---|---|---|---|---|---|---|---|---|
| ***# ECG*** | | 1,017,720 | 149,178 | 287,039 | 558,464 | 157,290 | 21,799 | 54,978 | 54,612 | 345,779 |
| **Avg. Exam per Patient** | | 5.52 (5.49 - 5.56) | 5.56 (5.45-5.67) | 5.51 (5.43-5.59) | 4.95 (4.90 - 4.99) | 4.93 (4.84-5.02) | 1.16 (1.15-1.16) | 1.08 (1.07-1.08) | 1.83 (1.83, 1.84) | 1.48 (1.48 - 1.48) |
| **Age** | | 60.40 (60.33 - 60.48) | 60.42 (60.23-60.61) | 60.47 (60.33-60.61) | 58.18 (58.07 - 58.30) | 57.81 (57.60-58.03) | 62.42 (61.95-62.89) | 64.57 (64.50-64.64) | 62.93 (62.81-63.05) | 51.00 (50.92 - 51.08) |
| ***# patients*** | | 184,210 | 26,835 | 52,098 | 112,902 | 31,906 | 18,869 | 51,055 | 29,427 | 233,770 |
| **Gender Distribution** | ***Male*** | 102,903 (55.86%) | 14,995 (55.88%) | 29,089 (55.84%) | 53,224 (47.14%) | 15,014 (47.06%) | 9,640 (51.09%) | 24,682 (48.34%) | 14,432 (49.04%) | 94,807 (40.56%) |
| | ***Female*** | 81,307 (44.14%) | 11,840 (44.12%) | 23,009 (44.16%) | 58,474 (51.79%) | 16,558 (51.90%) | 9,229 (48.91%) | 26,373 (51.66%) | 14,995 (50.95%) | 138,963 (59.44%) |

| | | | | | | | | | | |
|---|---|---|---|---|---|---|---|---|---|---|
| | ***Unknown*** | - | - | - | 1,204 (1.07%) | 334 (1.05%) | - | - | - | - |

**Supplemental Table 36. Data format for each dataset**

| **Dataset** | **Original Data Format** |
|---|---|
| MHI-ds | XML |
| MIMIC-IV | WFDB |
| Code-15 | HDF5 |
| UKB | XML |
| PTB | WFDB |
| CLSA | XML |
| JGH | DICOM |
| CSH | XML |
| MGH | XML |
| NYP | XML |
| UW | XML |
| UCSF | XML |

### References


1. Devlin, J., Chang, M.-W., Lee, K. & Toutanova, K. BERT: Pre-training of Deep Bidirectional Transformers for Language Understanding. (2019) doi:10.48550/arXiv.1810.04805.

2. Kligfield, P. *et al.* Recommendations for the Standardization and Interpretation of the Electrocardiogram. *J. Am. Coll. Cardiol.* **49**, 1109–1127 (2007).

3. Lin, T.-Y., Goyal, P., Girshick, R., He, K. & Dollár, P. Focal Loss for Dense Object Detection. (2017) doi:10.48550/ARXIV.1708.02002.

4. Loshchilov, I. & Hutter, F. Decoupled Weight Decay Regularization. (2017) doi:10.48550/ARXIV.1711.05101.

5. Heideman, M., Johnson, D. & Burrus, C. Gauss and the history of the fast fourier transform. *IEEE ASSP Mag* **1**, 14–21 (1984).

6. Cleveland, W. S. Robust Locally Weighted Regression and Smoothing Scatterplots. *J. Am. Stat. Assoc.* **74**, 829–836 (1979).

7. Biewald, L. Experiment Tracking with Weights and Biases. (2020).

8. Dosovitskiy, A. *et al.* An Image is Worth 16x16 Words: Transformers for Image Recognition at Scale. (2020) doi:10.48550/ARXIV.2010.11929.

9. Chen, C.-F., Fan, Q. & Panda, R. CrossViT: Cross-Attention Multi-Scale Vision Transformer for Image Classification. (2021) doi:10.48550/ARXIV.2103.14899.

10. Tan, M. & Le, Q. V. EfficientNet: Rethinking Model Scaling for Convolutional Neural Networks. (2019) doi:10.48550/ARXIV.1905.11946.

11. Tan, M. & Le, Q. V. EfficientNetV2: Smaller Models and Faster Training. (2021) doi:10.48550/ARXIV.2104.00298.

12. Gu, A. & Dao, T. Mamba: Linear-Time Sequence Modeling with Selective State Spaces. (2023) doi:10.48550/ARXIV.2312.00752.

13. He, K., Zhang, X., Ren, S. & Sun, J. Deep Residual Learning for Image Recognition. (2015) doi:10.48550/ARXIV.1512.03385.

14. Xie, S., Girshick, R., Dollár, P., Tu, Z. & He, K. Aggregated Residual Transformations for Deep Neural Networks. (2016) doi:10.48550/ARXIV.1611.05431.

15. Szegedy, C., Vanhoucke, V., Ioffe, S., Shlens, J. & Wojna, Z. Rethinking the Inception Architecture for Computer Vision. (2015) doi:10.48550/ARXIV.1512.00567.

16. Huang, G., Liu, Z., van der Maaten, L. & Weinberger, K. Q. Densely Connected Convolutional Networks. (2016) doi:10.48550/ARXIV.1608.06993.

17. Saito, T. & Rehmsmeier, M. The Precision-Recall Plot Is More Informative than the ROC Plot When Evaluating Binary Classifiers on Imbalanced Datasets. *PLoS ONE* **10**, e0118432 (2015).

18. Pedregosa, F. *et al.* Scikit-learn: Machine Learning in Python. *J. Mach. Learn. Res.* **12**, 2825–2830 (2011).

19. Iwana, B. K. & Uchida, S. Time Series Data Augmentation for Neural Networks by Time Warping with a Discriminative Teacher. (2020) doi:10.48550/ARXIV.2004.08780.

20. Lam, S. K., Pitrou, A. & Seibert, S. Numba: a LLVM-based Python JIT compiler. in *Proceedings of the Second Workshop on the LLVM Compiler Infrastructure in HPC* 1–6 (ACM, Austin Texas, 2015). doi:10.1145/2833157.2833162.

21. Kobayashi, T. Two-Way Multi-Label Loss. in *Proceedings of the IEEE/CVF Conference on Computer Vision and Pattern Recognition (CVPR)* 7476–7485 (2023).

22. Zhang, Y. *et al.* Simple and Robust Loss Design for Multi-Label Learning with Missing Labels. (2021) doi:10.48550/ARXIV.2112.07368.

23. Ben-Baruch, E. *et al.* Asymmetric Loss For Multi-Label Classification. (2020) doi:10.48550/ARXIV.2009.14119.

24. Agarap, A. F. Deep Learning using Rectified Linear Units (ReLU). (2018) doi:10.48550/ARXIV.1803.08375.

25. Xu, B., Wang, N., Chen, T. & Li, M. Empirical Evaluation of Rectified Activations in Convolutional Network. (2015) doi:10.48550/ARXIV.1505.00853.

26. Hendrycks, D. & Gimpel, K. Gaussian Error Linear Units (GELUs). (2016) doi:10.48550/ARXIV.1606.08415.

27. Klambauer, G., Unterthiner, T., Mayr, A. & Hochreiter, S. Self-Normalizing Neural Networks. (2017) doi:10.48550/ARXIV.1706.02515.

28. Misra, D. Mish: A Self Regularized Non-Monotonic Activation Function. (2019) doi:10.48550/ARXIV.1908.08681.

29. Ramachandran, P., Zoph, B. & Le, Q. V. Searching for Activation Functions. (2017) doi:10.48550/ARXIV.1710.05941.

30. Huang, G., Sun, Y., Liu, Z., Sedra, D. & Weinberger, K. Deep Networks with Stochastic Depth. (2016) doi:10.48550/ARXIV.1603.09382.

31. Srivastava, N., Hinton, G., Krizhevsky, A., Sutskever, I. & Salakhutdinov, R. Dropout: A Simple Way to Prevent Neural Networks from Overfitting. *J. Mach. Learn. Res.* **15**, 1929–1958 (2014).

32. Kingma, D. P. & Ba, J. Adam: A Method for Stochastic Optimization. (2014) doi:10.48550/ARXIV.1412.6980.

33. Liu, L. *et al.* On the Variance of the Adaptive Learning Rate and Beyond. (2019) doi:10.48550/ARXIV.1908.03265.

34. Ruder, S. An overview of gradient descent optimization algorithms. (2016) doi:10.48550/ARXIV.1609.04747.

35. Duchi, J., Hazan, E. & Singer, Y. Adaptive Subgradient Methods for Online Learning and Stochastic Optimization. *J. Mach. Learn. Res.* **12**, 2121–2159 (2011).

36. Kurbiel, T. & Khaleghian, S. Training of Deep Neural Networks based on Distance Measures using RMSProp. (2017) doi:10.48550/ARXIV.1708.01911.

37. Loshchilov, I. & Hutter, F. SGDR: Stochastic Gradient Descent with Warm Restarts. (2016) doi:10.48550/ARXIV.1608.03983.

38. Seetharaman, P., Wichern, G., Pardo, B. & Roux, J. L. AutoClip: Adaptive Gradient Clipping for Source Separation Networks. (2020) doi:10.48550/ARXIV.2007.14469.

39. Chen, T., Kornblith, S., Norouzi, M. & Hinton, G. A Simple Framework for Contrastive Learning of Visual Representations. (2020) doi:10.48550/ARXIV.2002.05709.

40. Grill, J.-B. *et al.* Bootstrap your own latent: A new approach to self-supervised Learning. (2020) doi:10.48550/ARXIV.2006.07733.

41. Assran, M. *et al.* Self-Supervised Learning from Images with a Joint-Embedding Predictive Architecture. (2023) doi:10.48550/ARXIV.2301.08243.

42. Oh, J., Chung, H., Kwon, J., Hong, D. & Choi, E. Lead-agnostic Self-supervised Learning for Local and Global Representations of Electrocardiogram. (2022) doi:10.48550/ARXIV.2203.06889.

43. Courty, B. *et al.* mlco2/codecarbon: v2.4.1. (2024) doi:10.5281/ZENODO.11171501.

44. ye, xiaoju. calflops: a FLOPs and Params calculate tool for neural networks in pytorch framework. (2023).

45. Environmental Protection Agency, E. Greenhouse Gas Emissions from a Typical Passenger Vehicle. (2023).

46. Ribeiro, M. T., Singh, S. & Guestrin, C. 'Why Should I Trust You?': Explaining the Predictions of Any Classifier. (2016) doi:10.48550/ARXIV.1602.04938.

47. Müller, R., Kornblith, S. & Hinton, G. When Does Label Smoothing Help? (2019) doi:10.48550/ARXIV.1906.02629.

48. Cubuk, E. D., Zoph, B., Shlens, J. & Le, Q. V. RandAugment: Practical automated data augmentation with a reduced search space. (2019) doi:10.48550/ARXIV.1909.13719.
